# Using multiple sampling strategies to estimate SARS-CoV-2 epidemiological parameters from genomic sequencing data

**DOI:** 10.1101/2022.02.04.22270165

**Authors:** Rhys P. D. Inward, Kris V. Parag, Nuno R. Faria

## Abstract

SARS-CoV-2 virus genomes are currently being sequenced at an unprecedented pace. The choice of viral sequences used in genetic and epidemiological analysis is important as it can induce biases that detract from the value of these rich datasets. This raises questions about how a set of sequences should be chosen for analysis, and which epidemiological parameters derived from genomic data are sensitive or robust to changes in sampling. We provide initial insights on these largely understudied problems using SARS-CoV-2 genomic sequences from Hong Kong, China, and the Amazonas State, Brazil. We consider sampling schemes that select sequences uniformly, in proportion or reciprocally with case incidence and which simply use all available sequences (unsampled). We apply *Birth-Death Skyline* and *Skygrowth* methods to estimate the time-varying reproduction number (*R*_*t*_) and growth rate (*r*_*t*_) under these strategies as well as related *R*_*0*_ and date of origin parameters. We compare these to estimates from case data derived from *EpiFilter*, which we use as a reference for assessing bias. We find that both *R*_*t*_ and *r*_*t*_ are sensitive to changes in sampling whilst *R*_*0*_ and the date of origin are relatively robust. Moreover, we find that analysis using unsampled datasets, which reflect an opportunistic sampling scheme, result in the most biased *R*_*t*_ and *r*_*t*_ estimates for both our Hong Kong and Amazonas case studies. We highlight that sampling strategy choices may be an influential yet neglected component of sequencing analysis pipelines. More targeted attempts at genomic surveillance and epidemic analyses, particularly in settings with limited sequencing capabilities, are necessary to maximise the informativeness of virus genomic datasets.

## INTRODUCTION

Severe acute respiratory syndrome coronavirus 2 (SARS-CoV-2) is an enveloped single-stranded zoonotic RNA virus belonging to the *Betacoronavirus* genus and *Coronaviridae* family^1^. It was first identified in late 2019 in a live food market in Wuhan City, Hubei Province, China^2^. Within a month, SARS-CoV-2 had disseminated globally through sustained human-to-human transmission. It was declared a public health emergency of international concern on the 30th of January 2020 by the World Health Organisation^3^. Those infected with SARS-CoV-2 have phenotypically diverse symptoms ranging from mild fever to multiple organ dysfunction syndromes and death^4^.

Despite the implementation of non-pharmaceutical interventions (NPIs) and rollout of vaccination programmes in many countries to control their epidemics, as of the 16^th^ July 2022, over 557 million SARS-CoV-2 cases and 6.3 million deaths have been reported worldwide^5^. These NPIs can vary within and between countries and include restrictions on international and local travel, school closures, social distancing measures and the isolation of infected individuals and their contacts^6^. The key aim of NPIs is to reduce epidemic transmission, often measured by epidemiological parameters such as the time-varying effective reproduction number (*R*_*t*_ at time *t*) and growth rate (*r*_*t*_), which both provide updating measures of the rate of spread of a pathogen (see Table 1 for detailed definitions)^7,8^.

**Table 1:** Key parameters and definitions for SARS-CoV-2

| Parameter | Definition |
| --- | --- |
| Basic reproduction number ($R_0$) | Average number of individuals infected by a single infected person in a fully susceptible population |
| Time-varying or effective reproduction number ($R_t$) | Average number of secondary infections generated per effective primary case at a certain time point and in the presence of susceptible depletion or interventions |
| Growth rate ($r_t$) | Rate of change of the logarithm of the number of new cases (i.e., the case incidence) per unit of time |
| Incubation period | Time between infection and symptom onset |
| Infectious period | Period in which an infectious host can transmit infectious agents to a susceptible individual |
| Generation interval | Time between infection events in an infector–infectee pair |
| Time of the most recent common ancestor or origin time | Date in which viral variant is thought to have emerged |
| Serial Interval | Time between symptom onsets in an infector–infectee pair |

However, there is currently great difficulty in estimating and comparing epidemiological parameters derived from case and death data globally due to disparities in molecular diagnostic surveillance and notification systems between countries. Further, even if data are directly comparable, the choice of epidemiological parameter can implicitly shape insights into how NPIs influence transmission potential^9,10^. As such, there is a need to supplement traditional estimates with information derived from alternative data sources, such as genomic data^11^, to gain improved and more robust insights into viral transmission dynamics^12,13^.

Phylodynamic analysis of virus genome sequences have increasingly been used for studying emerging infectious diseases, as seen during the current SARS-CoV-2 pandemic^14–17^, recent Ebola virus epidemics in Western Africa^18^ and the Zika epidemic in Brazil and the Americas^19,20^. Transmissibility parameters such as the basic reproduction number (*R*_*0*_), *R*_*t*_ and *r*_*t*_ can be directly inferred from genomic sequencing data or from epidemiological data, while other epidemiological parameters such as the time of the most recent common ancestor (TMRCA) of a given viral variant or lineage can only be estimated from genomic data. This is of particular importance for variants of concern (VOC), genetic variants with evidence of increased transmissibility, more severe disease, and/or immune evasion. VOC are typically detected through virus genome sequencing and only limited inferences can be made using epidemiological data alone^21^.

Currently, SARS-CoV-2 virus genomes from COVID-19 cases are being sequenced at an unprecedented pace providing a wealth of virus genomic datasets^22^. There are currently over 11.9 million genomic sequences available on GISAID, an open-source repository for influenza and SARS-CoV-2 genomic sequences^23^. These rich datasets can be used to provide an independent perspective on pathogen dynamics and can help validate or challenge parameters derived from epidemiological data. Specifically, the genomic data can potentially overcome some of the limitations and biases that can result from using epidemiological data alone. For example, genomic data are less susceptible to changes at the government level such as alterations to the definition of a confirmed case and changes to notification systems^24,25^. Inferences from virus genomic data improve our understanding of underlying epidemic spread and can facilitate better-informed infection control decisions^26^. However, these advantages are not straightforward to realise. The added value of genomic data depends on two related variables: sampling strategy and computational complexity.

The most popular approaches used to investigate changes in virus population dynamics include the Bayesian Skyline Plot^27^ and Skygrid^28^ models and the Birth-Death Skyline (BDSKY)^29^. These integrate Markov Chain Monte Carlo (MCMC) procedures and often converge slowly on large datasets^30^. As such, currently available SARS-CoV-2 datasets containing thousands of sequences become computationally impractical to analyse and sub-sampling is necessary. Although previous studies have examined how sampling choices might influence phylodynamic inferences^30–34^, this remains a neglected area of study^35^, particularly concerning SARS-CoV-2 for which sequencing efforts have been unprecedented ^36^. To our knowledge, there are no published studies concerning SARS-CoV-2 which explore the effect that sampling strategies can have on the phylodynamic reconstruction of key transmission parameters. Incorrectly implementing a sampling scheme or ignoring its importance can mislead inferences and introduce biases^30,37^.

This raises the important question that motivates our analysis: how should sequences be selected for phylodynamic analysis and which parameters are sensitive or robust to changes in different sampling schemes. Here we explore how diverse sampling strategies in genomic sequencing may affect the estimation of key epidemiological parameters. We estimate *R*_*0*_, *R*_*t*_, and *r*_*t*_ from genomic sequencing data under different sampling strategies from a location with higher genomic coverage represented by Hong Kong, and a location with lower genomic coverage represented by the Amazonas state, Brazil. We then compare our estimates against those derived from reference case data. By benchmarking genomic inferences against those from case data we can better understand the impact that sampling strategies may have on phylodynamic inference, bolster confidence in estimates of genomic-specific parameters such as the origin time (or TMRCA) and improve the interpretation of estimates from areas with heterogeneous genomic coverage.

## METHODS

### Empirical Estimation of the Reproduction Number, Time-varying Effective Reproduction Number, and Growth Rate

#### Epidemiological Datasets

Two sources of data from the Amazonas state, Brazil, and one source of data from Hong Kong were used to calculate empirical epidemiological parameters. For the Amazonas state, case data from the SIVEP-Gripe (Sistema de Informação de Vigilância Epidemiológica da Gripe) SARI (severe acute respiratory infections) database from the 30^th^ of November 2020 up to 7^th^ of February 2021 were used. Here we were interested in cases caused by the Gamma VOC first detected in Manaus^14^. The number of Gamma cases was calculated by using the proportion of Gamma viral sequences uploaded to GISAID within each week (Supplementary Figure 1). For Hong Kong, all case data were extracted from the Centre of Health Protection, Department of Health, the Government of the Hong Kong Special Administrative region up to the 7^th^ of May 2020. Due to lags in the development of detectable viral loads, symptom onset and subsequent testing^38^; the date on which each case was recorded was left shifted by 5 days within our models^39^ to account for these delays in both datasets.

#### Basic Reproduction Number

The *R*_*0*_ parameter was estimated using a time series of confirmed SARS-CoV-2 cases from both Hong Kong and the Amazonas state. To avoid the impact of NPIs, only data up to the banning of mass gathering in Hong Kong (27^th^ March 2020) and until the imposition of strict restrictions in the Amazonas state (12^th^ January 2021) were used. We estimated *R*_*0*_ from weekly counts of confirmed cases using maximum likelihood methods. The weekly case counts were assumed to be Poisson distributed and were fitted to a closed Susceptible-Exposed-Infectious-Recovered (SEIR) model (equation (1)) by maximising the likelihood of observing the data given the model parameters (Table 2). Subsequently, the log-likelihood was used to calculate the R_0_ by fitting β, the effective contact rate.

**Table 2:** This shows the parameter estimates used within the deterministic SEIR model.

| Parameter | Description | Value (source) |
| --- | --- | --- |
| $R_0 = \beta\alpha$ | Basic Reproduction Number | Estimated |
| N | Population of Hong Kong | 7,481,800 persons<sup>40</sup> |
|  | Population of Amazonas state | 4,207,714 persons<sup>41</sup> |
| $\beta$ | Effective Contact Rate | Estimated |
| $\alpha$ | Infectious Period | 0.07 day<sup>-1</sup> <sup>42</sup> |
| $\lambda$ | Force of Infection | Estimated |
| $\gamma$ | Progression from E to I | 5.26 day<sup>-1</sup> <sup>43</sup> |
| $\sigma$ | Progression from I to R | 14.3 day<sup>-1</sup> <sup>42</sup> |
| S | Estimated number of Susceptibles | Estimated |
| E | Estimated number of Exposed | Estimated |
| I | Number of Infected | Weekly case counts |
| R | Estimated number of Recovered | Estimated |

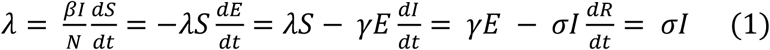

To generate approximate 95% confidence intervals (CIs) for *R*_*0*_, non-parametric bootstrapping was used with 1000 iterations.

#### Time-varying Effective Reproduction Number

To estimate *R*_*t*_ from case time series data the *EpiFilter* method^44^ was used. *EpiFilter* describes transmission using a renewal model; a general and popular framework that can be applied to infer the dynamics of numerous infectious diseases from case incidence^45^. This model describes how the number of new cases (incidence) at time *t* depends on *R*_*t*_ at that specified time point and the past incidence, which is summarised by the cumulative number of cases up to each time point weighted by the generation time distribution. *EpiFilter* integrates both Bayesian forward and backward recursive smoothing. This improves *R*_*t*_ estimates by leveraging the benefits of two of the most popular *R*_*t*_ estimation approaches: *EpiEstim* ^46^ and the Wallinga-Teunis method^47^. *EpiFilter* minimises the mean squared error in estimation and reduces dependence on prior assumptions, making it a suitable candidate for deriving reference estimates. We use these to benchmark estimates independently obtained from genomic data. We assume the generation time distribution is well approximated by the serial interval (SI) distribution^46^, which describes the times between symptom onsets between an infector–infectee pair. We describe the specific SI distributions that we used in the next subsection.

#### Growth Rate

After *R*_*t*_ has been inferred, the Wallinga-Lipsitch equation for a gamma distributed generation time distribution (equation (2)) was used to estimate the exponential epidemic *r*_*t*_ ^48^. The SI for Hong Kong was derived from a systematic review and meta-analysis^49^ and a study exploring SI in Brazil was used for the Amazonas datasets^50^. The SI was assumed to be gamma distributed. The gamma distribution is represented by gamma (ε, γ) with ε *and* γ being the shape and scale parameters respectively.

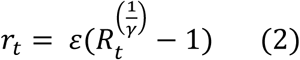

### SARS-CoV-2 Brazilian Gamma VOC and Hong Kong datasets

All high-quality (<1% N, or non-identified nucleotide), complete (>29 kb) SARS-CoV-2 genomes were downloaded from GISAID^23^ for Hong Kong (up to 7^th^ May 2020) and the Amazonas state, Brazil (from 30th November 2020 up to 7^th^ February 2021). Using the Accession ID of each sequence, all sequences were screened and only sequences previously analysed and published in PubMed, MedRxiv, BioRxiv, virological or Preprint repositories were selected for subsequent analysis. For both datasets, sequence alignment was conducted using MAFFTV.7^51^. The first 130 base pairs (bp) and last 50 bps of the aligned sequences were trimmed to remove potential sequencing artefacts in line with the Nextstrain protocol^52^. Both datasets were then processed using the Nextclade pipeline for quality control (https://clades.nextstrain.org/). Briefly, the Nextclade pipeline examines the completeness, divergence, and ambiguity of bases in each genetic sequence. Only sequences deemed ‘good’ by the Nextclade pipeline were selected. Subsequently, all sequences were screened for identity and in the case of identical sequences, for those with the same location, collection date, only one such isolate was used. Moreover, PANGO lineage classification was conducted using the Pangolin^22^ (v2.3.3) software tool (http://pangolin.cog-uk.io) on sequences from the Amazonas state and only those with the designated P.1/Gamma lineage were selected (Supplementary Figure 1).

### Maximum Likelihood tree reconstruction

Maximum likelihood phylogenetic trees were reconstructed using IQTREE2^53^ for both datasets. A TIM2 model of nucleotide substitution with empirical base frequencies and a proportion of invariant sites was used as selected for by the ModelFinder application^54^ for the Hong Kong dataset. For the Brazilian dataset, a TN model of nucleotide substitution^55^ with empirical base frequencies was selected for. To assess branch support, the approximate likelihood-ratio test based on the Shimodaira–Hasegawa-like procedure with 1,000 replicates^56^, was used.

### Root-to-tip regression

To explore the temporal structure of both the Brazilian and Hong Kong dataset, TempEst v.1.5.3^57^ was used to regress the root-to-tip genetic distances against sampling dates (yyyy-mm-dd). The ‘best-fitting’ root for the phylogeny was found by maximising the R^2^ value of the root-to-tip regression (Supplementary Figure 2). Several sequences showed incongruent genetic diversity and were discarded from subsequent analyses. This resulted in a final dataset of N = 117 Hong Kong sequences and N = 196 Brazilian sequences. The gradient of the slopes (clock rates) provided by TempEst were used to inform the clock prior in the phylodynamic analysis.

### Subsampling for analysis

Four retrospective sampling schemes were used to select a subsample of Amazonas and Hong Kong sequences. Each sampling period was broken up into weeks with each week being used as an interval according to a temporal sampling scheme (without replacement). This temporal sampling scheme was based on the number of reported cases of SARS-CoV-2.

The temporal sampling schemes that we explored were:

- **No sampling strategy applied:** All sequences were included without a sampling strategy applied (equivalent to opportunistic sampling).
- **Proportional sampling**: Weeks are chosen with a probability proportional to the value of the number of incident cases in each epi-week.
- **Uniform sampling:** All weeks have equal probability.
- **Reciprocal-proportional sampling:** Weeks are chosen with a probability proportional to the reciprocal of the incident number of cases in each epi-week.

These sampling schemes were inspired by those recommended by the WHO for practical use in different settings and scenarios^58^. Proportional sampling is equivalent to representative sampling, uniform sampling is equivalent to fixed sampling whilst the unsampled data includes all sampled sequences. Reciprocal-proportional sampling is not commonly applied in practice and was used as a control within this study.

### Bayesian Evolutionary Analysis

Date molecular clock phylogenies were inferred for all sampling strategies applied to the Amazonas and Hong Kong dataset using BEAST v1.10.4^59^ with BEAGLE library v3.1.0^60^ for accelerated likelihood evaluation. For both the Amazonas and Hong Kong datasets, a HKY substitution model with gamma-distributed rate variation among sites and four rate categories was used to account for among-site rate variation^61^. A strict clock molecular clock model was chosen. Both the Amazonas and Hong Kong dataset were analysed under a flexible non-parametric skygrid tree prior^62^. Four independent MCMC chains were run for both datasets.

For the Amazonas dataset, each MCMC chain consisted of 250,000,000 steps with sampling every 50,000 steps. Meanwhile, for the Hong Kong dataset, each MCMC chain consisted of 200,000,000 steps with sampling every 40,000 steps. For both datasets, the four independent MCMC runs were combined using LogCombiner v1.10.4^59^. Subsequently, 10% of all trees were discarded as burn in, and the effective sample size of parameter estimates were evaluated using TRACER v1.7.2^63^. An effective sample size of over 200 was obtained for all parameters. Maximum clade credibility (MCC) trees were summarised using Tree Annotator^59^.

### Phylodynamic Reconstruction

#### Estimation of the Basic and Time-varying Effective Reproduction Numbers

The Bayesian birth-death skyline (BDSKY) model^29^ implemented within BEAST 2 v2.6.5^64^ was applied to estimate the time-varying transmissibility parameter *R*_*t*_ (Table 3). An HKY substitution model with a gamma-distributed rate variation among sites and four rate categories^61^ was used alongside a strict molecular clock model. The selected number of intervals for both datasets was 5, representing *R*_*t*_ changing every 2.5 weeks for the Hong Kong datasets and every 2 weeks for the Brazilian datasets, with equidistant intervals per step. An exponential distribution was used with a mean of 36.5y^-1^ for the rate of becoming infectious, assuming a mean duration of infection of 10 days^15^. A uniform distribution prior was used for the sampling proportion, which models changes in case ascertainment. Four independent MCMC chains were run for 50 million MCMC steps with sampling every 5000 steps for each dataset. These MCMC runs were combined using LogCombiner v2.6.5.^64^ and the effective sample size of parameter estimates evaluated using TRACER v1.7.2^63^. We obtained an effective sample size above 200 for all parameters (indicating convergence) and plotted all results using the bdskytools R package (https://github.com/laduplessis/bdskytools).

**Table 3:**
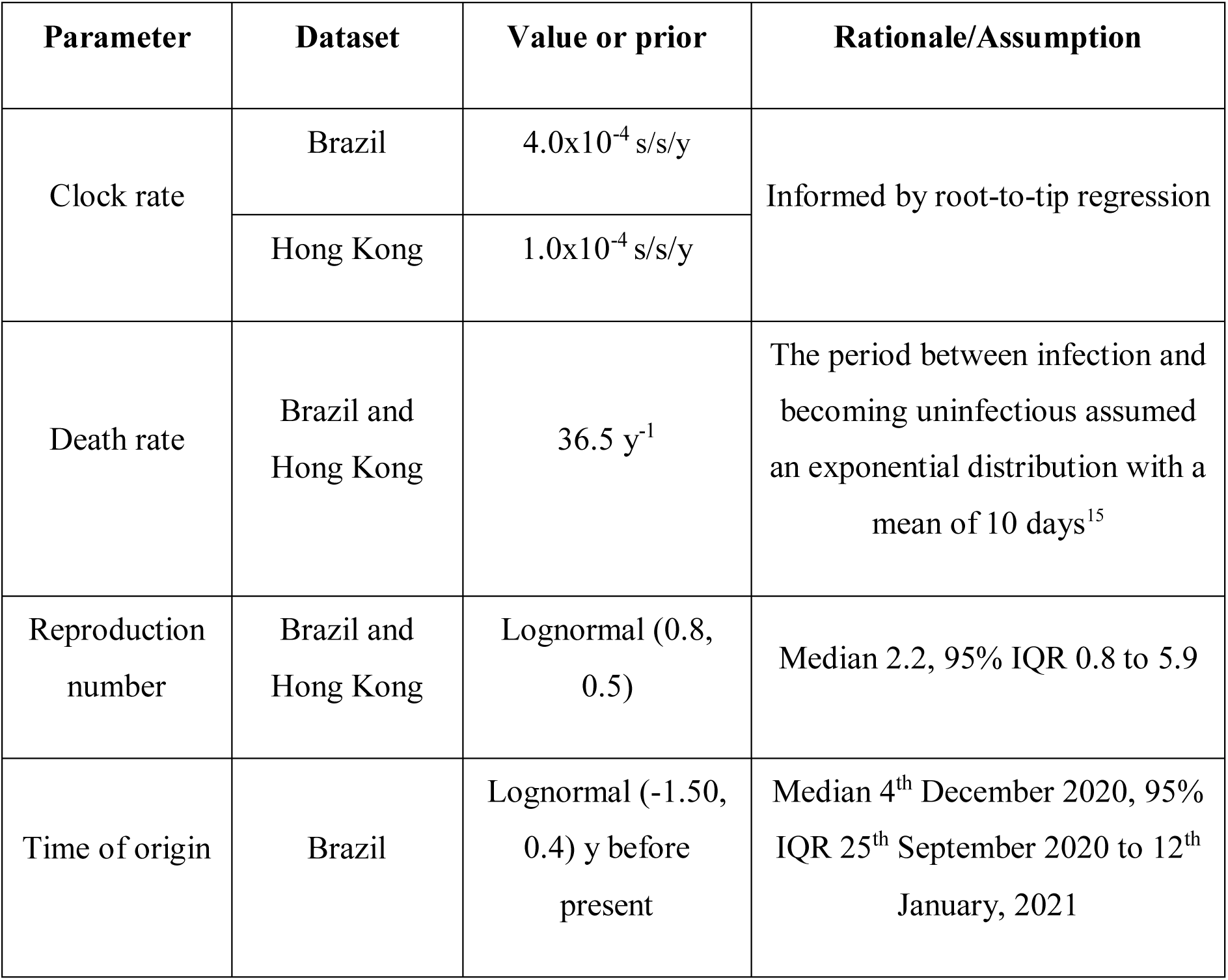
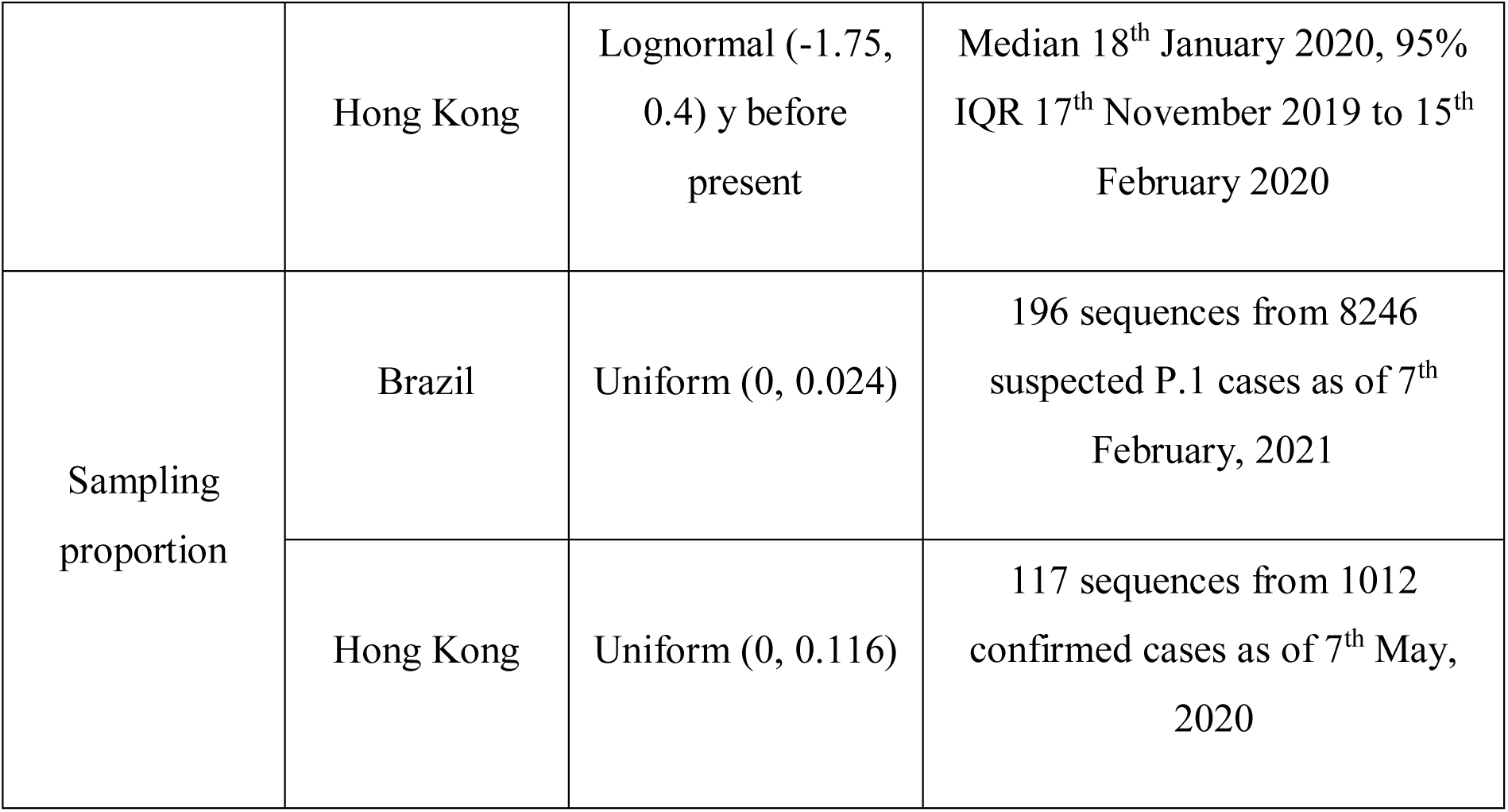
Values and priors for the parameters of the BDSKY model. s/s/y=substitutions per site per year.

| Parameter | Dataset | Value or prior | Rationale/Assumption |
| --- | --- | --- | --- |
| Clock rate | Brazil | $4.0 \times 10^{-4}$ s/s/y | Informed by root-to-tip regression |
| | Hong Kong | $1.0 \times 10^{-4}$ s/s/y | |
| Death rate | Brazil and Hong Kong | $36.5 \text{ y}^{-1}$ | The period between infection and becoming uninfected assumed an exponential distribution with a mean of 10 days<sup>15</sup> |
| Reproduction number | Brazil and Hong Kong | Lognormal (0.8, 0.5) | Median 2.2, 95% IQR 0.8 to 5.9 |
| Time of origin | Brazil | Lognormal (-1.50, 0.4) y before present | Median 4<sup>th</sup> December 2020, 95% IQR 25<sup>th</sup> September 2020 to 12<sup>th</sup> January, 2021 |
|  | Hong Kong | Lognormal (-1.75, 0.4) y before present | Median 18<sup>th</sup> January 2020, 95% IQR 17<sup>th</sup> November 2019 to 15<sup>th</sup> February 2020 |
| Sampling proportion | Brazil | Uniform (0, 0.024) | 196 sequences from 8246 suspected P.1 cases as of 7<sup>th</sup> February, 2021 |
|  | Hong Kong | Uniform (0, 0.116) | 117 sequences from 1012 confirmed cases as of 7<sup>th</sup> May, 2020 |

#### Estimation of Growth Rates

For each dataset, a scaled proxy for *r*_*t*_ was obtained from the *Skygrowth* method^65^ within R. *Skygrowth* uses a non-parametric Bayesian approach to apply a first-order autoregressive stochastic process on the growth rate of the effective population size. The MCMC chains were run for one million iterations for each dataset on their MCC tree with an Exponential (10^−5^) prior on the smoothing parameter. The *Skygrowth* model was parameterised assuming that the effective population size of SARS-COV-2 could change every two weeks. To facilitate a comparison of the scaled proxy for *r*_*t*_ estimated by *Skygrowth* and exponential *r*_*t*_ estimated by *EpiFilter*, the *r*_*t*_ estimated by the *Skygrowth* method was rescaled to the exponential growth rate. This was achieved by adding a gamma rate variable to the scaled proxy for *r*_*t*_, which assumed a mean duration of infection of 10 days^15^, to calculate *R*_*t*_. Subsequently, the Wallinga-Lipsitch equation (Equation 2) was used to convert *R*_*t*_ into the exponential growth rate^48^.

### Comparing Parameter Estimates from Genetic and Epidemiological Data

To compare estimates derived from epidemiological and genetic data the Jensen-Shannon divergence (D_JS_)^66^, which measures the similarity between two probability mass functions (PMFs), was applied. The D_JS_ offers a formal information theoretic evaluation of distributions and is more robust than comparing Bayesian credible intervals (BCIs) since it considers both the shape and spread of a given distribution. The D_JS_ is a symmetric and smoothed version of the Kullback-Leibler divergence (D_KL_) and is commonly used in the fields of machine learning and bioinformatics. The D_KL_ between two PMFs, P and Q, is defined in equation (3) below^67^, with *x* spanning the common domain of those PMFs.

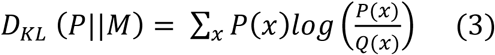

To calculate the PMF for each epidemiological parameter, the cumulative probability density function was extracted for each model, converted to a probability density function and a discretisation procedure was applied to generate the associated PMF.

The Jensen-Shannon distance (JSD) is a metric which takes the square-root of the total D_JS_ and is the metric that we used to compare parameter estimations from differing sampling strategies. The JSD can be calculated using Equation 4 with P and Q representing the two probability distributions and D_KL_ as the KL divergence. A smaller JSD metric indicates that two probability distributions (P and Q) are more similar with a Jensen-Shannon distance of 0 uniquely indicating that both distributions are equivalent. The mean JSD was taken over all intervals for the BDSKY and *Skygrowth* models to obtain an overall measure of the level of estimated similarity across the epidemic trajectory. We do not expect the JSD to perfectly align with the 95% highest posterior density interval if the shapes of distributions from different schemes are very different.

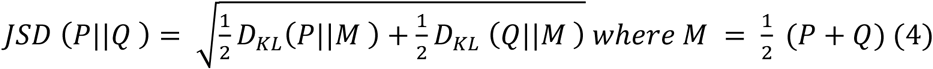

### Data availability

Code and data for reproducing the analyses presented in this study are freely available at https://github.com/rhysinward/Phylodynamic-Subsampling.

## RESULTS

### Sampling Schemes

#### Hong Kong

Hong Kong reacted rapidly upon learning of the emergence of SARS-CoV-2 in Wuhan, Hubei province, China, by declaring a state of emergency on the 25th of January 2020 and by mobilising intensive surveillance schemes in response to initial cases^68^. This appeared to be successful in controlling the first wave of cases. However, due to imported cases from Europe and North America, a second wave of SARS-CoV-2 infections emerged prompting stricter NPIs such as the closure of borders and restrictions on gatherings ^68^. Following these measures, the incidence of SARS-CoV-2 rapidly decreased (Figure 1). Hong Kong has a high sampling intensity with 11.6% of confirmed cases sequenced during our study period. Further, Hong Kong has high quality case data with a high testing rate through effective tracing of close contacts, testing of all asymptomatic arriving travellers and all patients with pneumonia^69^.

**Figure 1.**
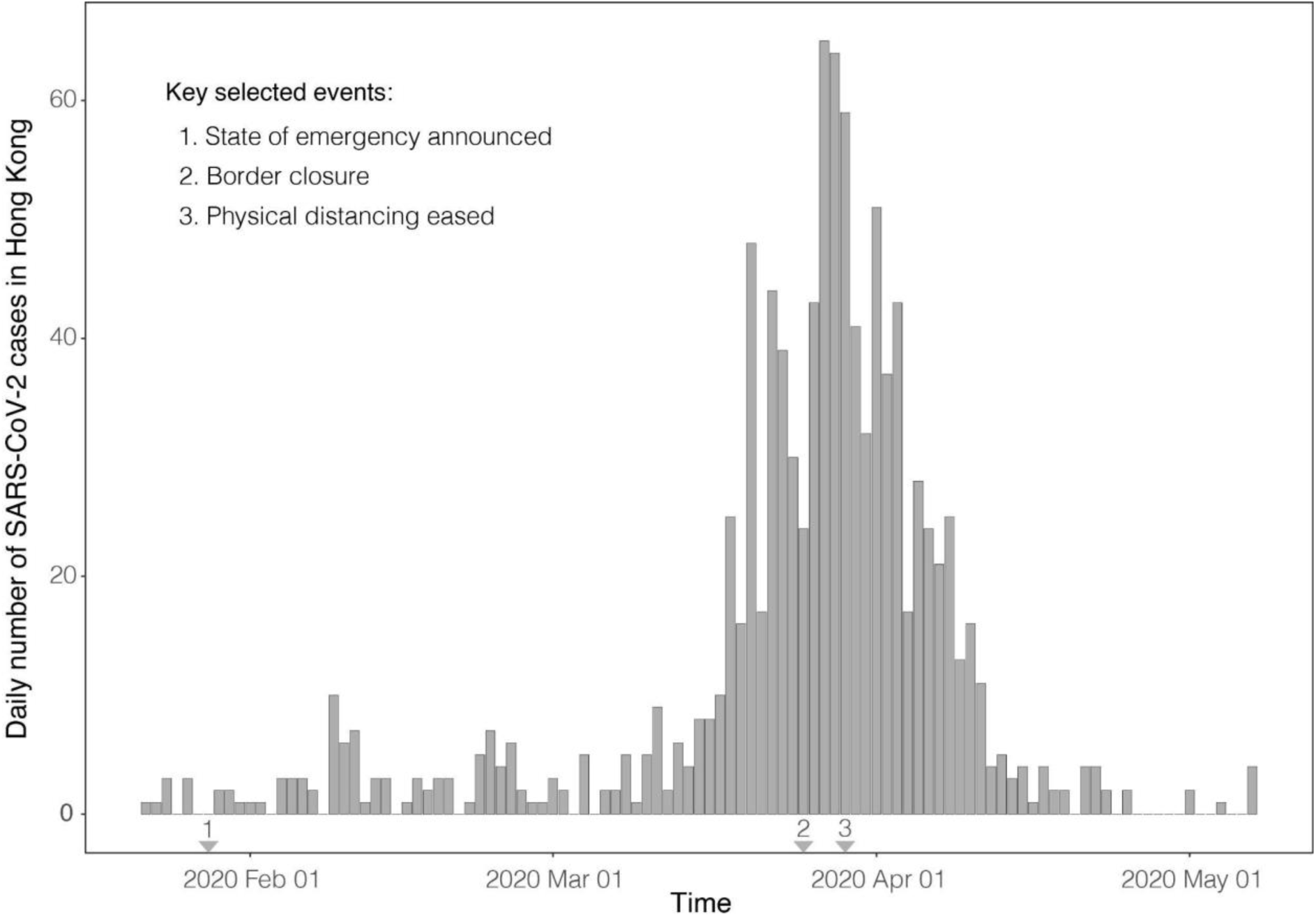
Confirmed incident SARS-CoV-2 cases from Hong Kong until 7^th^ of May 2020. The arrows represent policy change-times^68^.

The number of cases within Hong Kong for each week was used to inform the sampling schemes used within this study. This resulted in the unsampled scheme having N = 117 sequences, the proportional sampling scheme having N = 54 sequences, the uniform sampling scheme having N = 79 and the reciprocal-proportional sampling scheme having N = 84 sequences (Supplementary Figure 3).

#### Amazonas

The Amazonas state of Brazil had its first laboratory confirmed case of SARS-CoV-2 in March 2020 in a traveller returning from Europe^70^. After a first large wave of SARS-CoV-2 infections within the state that peaked in early May 2020 (Figure 2), the epidemic waned, cases dropped, remaining stable until mid-December 2020. The number of cases then started growing exponentially, ushering in a second epidemic wave. This second wave peaked in January 2021 (Figure 2) and coincided with the emergence of a new SARS-CoV-2 VOC, designated P.1/Gamma^14^.

**Figure 2.**
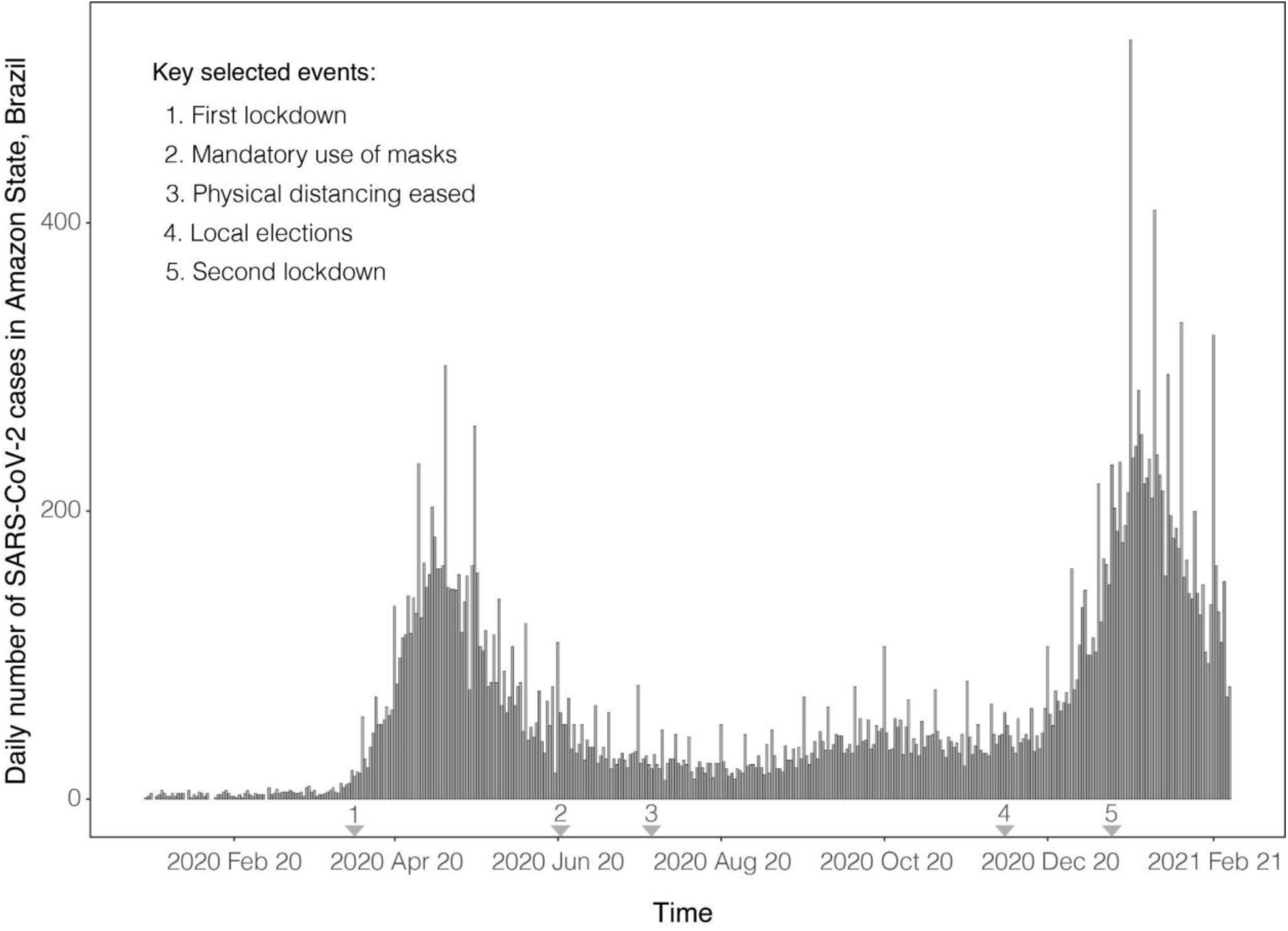
Confirmed incident SARS-CoV-2 cases from Amazonas state, north Brazil until 7^th^ of February 2021. The arrows represent key policy change-times^72^.

To combat this second wave, the Government of the Amazonas state suspended all non-essential commercial activities on the 23rd of December 2020 (http://www.pge.am.gov.br/legislacao-covid-19/). However, in response to protests, these restrictions were reversed, and cases continued to climb. On the 12th of January, when local transmission of P.1/Gamma was confirmed in Manaus, capital of Amazonas state^71^, NPIs were re-introduced (http://www.pge.am.gov.br/legislacao-covid-19/) which seemed to be successful in reducing the case incidence in the state. However, cases remained comparatively high (Figure 4). Amazonas has a sampling intensity with 2.4% of suspected P.1/gamma cases sequenced during our study period.

The number of cases within the Amazonas state informed the sampling schemes used within this study. This resulted in the unsampled scheme having N = 196 sequences, the proportional sampling scheme having N = 168 sequences, the uniform sampling scheme having N = 150 and the reciprocal-proportional sampling scheme having N = 67 sequences (Supplementary Figure 4).

### Root-to-tip Regression

The correlation (R^2^) between genetic divergence and sampling dates for the Hong Kong datasets ranged between 0.36 and 0.52 and between 0.13 and 0.20 for the Amazonas datasets (Supplementary Figure 2). This implies that the Hong Kong datasets have a stronger temporal signal. This is likely due to the Hong Kong datasets having a wider sampling interval (106 days) compared to the Amazonas datasets (69 days). A wider sampling interval can lead to a stronger temporal signal^73^. The gradient (rate) of the regression ranged from 1.16×10^−3^ to 2.09×10^−3^ substitutions per site per year (s/s/y) for the Hong Kong datasets and 4.41×10^−4^ to 5.30×10^−4^ s/s/y for the Amazonas datasets.

### Estimation of Evolutionary Parameters

The mean substitution rate (measured in units of number of s/s/y) and the TMRCA was estimated in BEAST, for both datasets, and the estimation from all sampling schemes was compared.

#### Hong Kong

For Hong Kong, the mean substitution rate per site per year ranged from 9.16×10^−4^ to 2.09×10^−3^ with sampling schemes all having overlapping Bayesian credible intervals (BCIs) (Supplementary table 2; Supplementary Figure 5A). This indicates that the sampling scheme did not have a significant impact on the estimation of the clock rate. Moreover, the clock rate is comparable to estimations from the root-to-tip regression and to early estimations of the mean substitution rate per site per year of SARS-CoV-2 (Duchene et al., 2020).

Molecular clock dating of the Hong Kong dataset indicates that the estimated time of the most common recent ancestor was around December 2020 (Figure 3B; Supplementary Table 2). This is a few weeks before the first confirmed case which was reported on the 18th of January 2021. Once again, all sampling strategies have overlapped BCIs and with the range in means differing by around three weeks, a relatively short time scale, suggesting that the sampling scheme does not significantly impact the estimation of the TMRCA.

**Figure 3.**
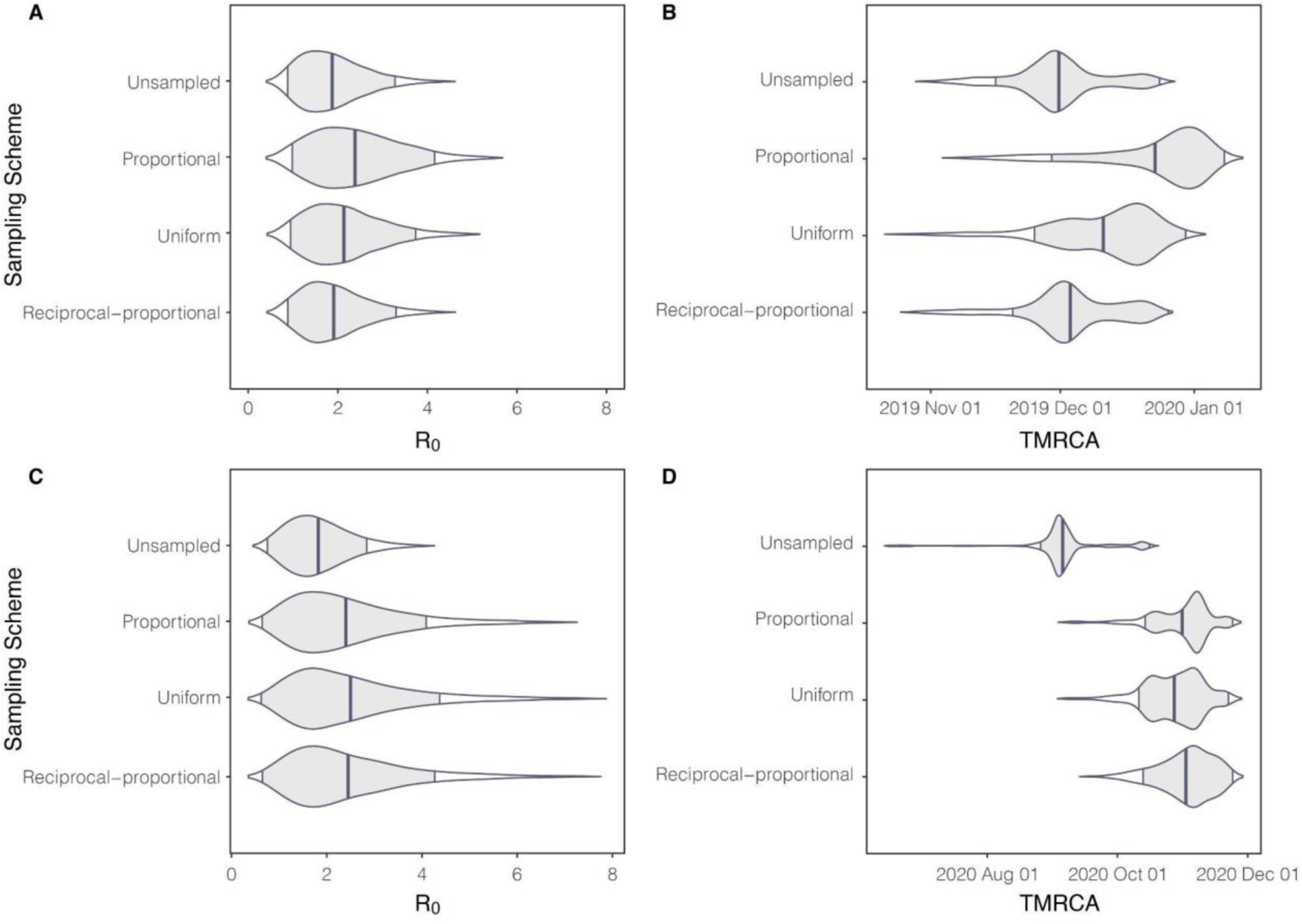
*R*_*0*_ estimated from BDSKY (using sequence data) and TMRCA for Hong Kong and Brazil. Figure 1A and B represent Hong Kong and Figure 1C and D represent the Amazonas. The central line represents the posterior mean and with intervals representing 95% highest posterior density interval.

#### Brazil

For the Gamma VOC in the Amazonas state, the mean substitution rate ranged from 4.00×10^−4^ to 5.56×10^−4^ s/s/y with all sampling schemes having overlapped BCIs (Figure 3D, Supplementary Table 2; Supplementary Figure 5B). This indicates that sampling strategy does not impact the estimation of the clock rate, supporting findings from the Hong Kong dataset. This also supports estimations from the root-to-tip analysis (Supplementary Figure 2).

Molecular clock dating estimated a TMRCA mean around late October to early November (Figure 3D; Supplementary Table 2). This is around five weeks before the date of the first P.1 case identified in Manaus used in our study. All sampling schemes have overlapping BCI consistent with the conclusion from the Hong Kong data that TMRCA is relatively robust to sampling.

### Estimation of Basic Reproduction Number

We found from using genomic data, Hong Kong had a posterior mean *R*_*0*_ estimate of 2.07 (Figure 3A) across all sampling strategies. Using a proportional sampling strategy gave the highest posterior mean *R*_*0*_ estimate of 2.38 with the unsampled sampling strategy giving the lowest posterior mean *R*_*0*_ estimate of 1.87. Overall, Brazil had a higher posterior mean *R*_*0*_ estimate with a value of 2.24 (Figure 3B) across all sampling strategies. The uniform sampling strategy yielded the highest posterior mean *R*_*0*_ estimate of 2.50 while the unsampled sampling strategy gave the lowest one of 1.82. Using case data, we found similarly found that Hong Kong had a lower *R*_*0*_ of 2.17 (95% credible interval (CI) = 1.43 - 2.83) when compared to Amazonas which had a *R*_*0*_ of 3.67 (95% CI = 2.83 – 4.48). All sampling schemes for both datasets were characterised by similar *R*_*0*_ values (Figure 3) indicating that the estimation of *R*_*0*_ is robust to changes in sampling scheme.

### Time-varying Reproduction number and Growth rate

We estimate *R*_*t*_ and *r*_*t*_ for local SARS-CoV-2 epidemics in Hong Kong and Amazonas, Brazil. Our main results showing these two parameters and JSD metrics are shown in Figures 4-8.

**Figure 4:**
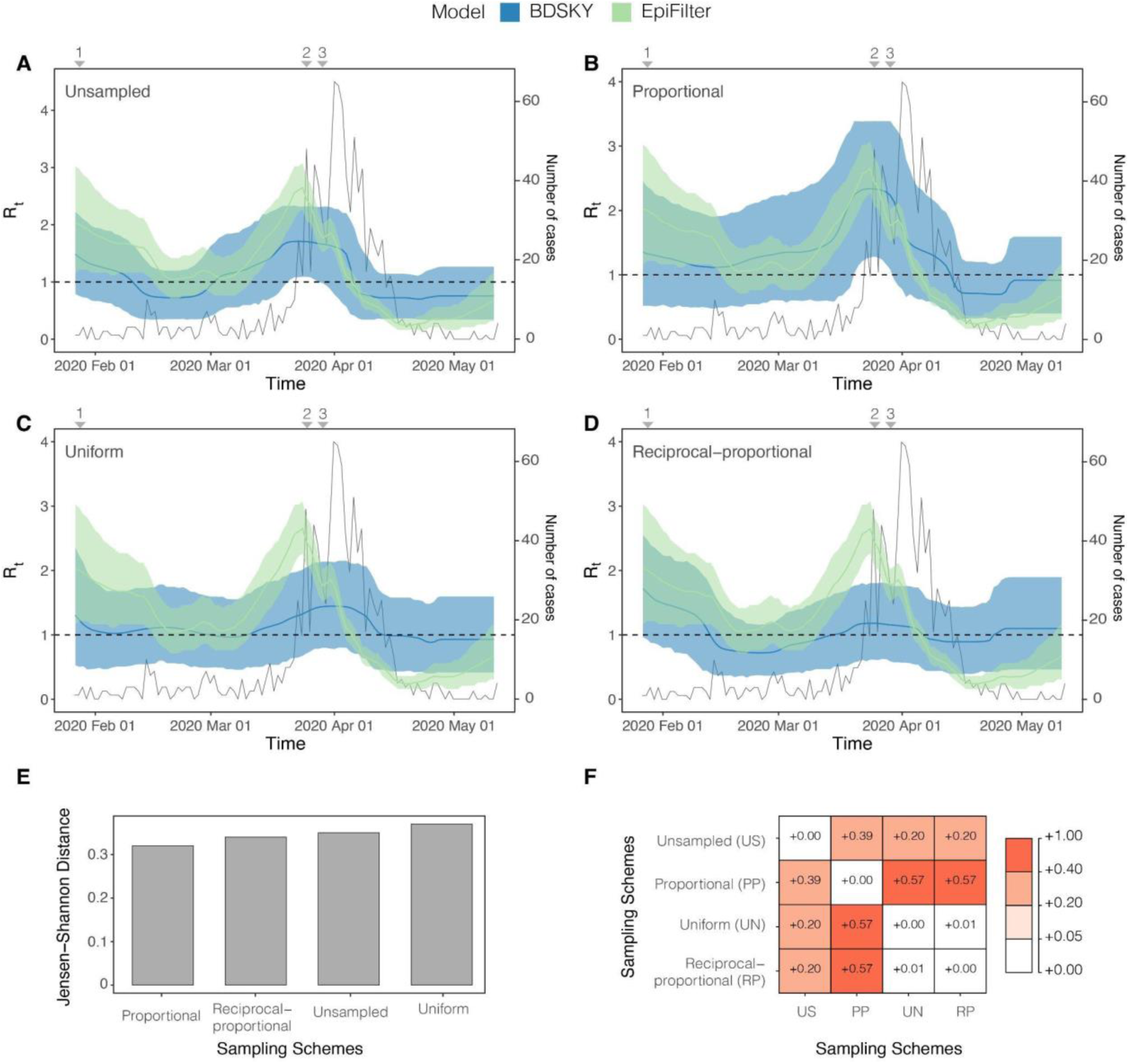
*R*_*t*_ estimated from both the BDSKY and *EpiFilter* methods for Hong Kong. Titles indicate the sampling scheme used in panels A-D. The light-shaded area represents the 95% highest posterior density interval. The solid line represents the mean *R*_*t*_ estimate with *EpiFilter* in green and BDSKY in blue. The black line plots the number of cases. We refer to Figure 1 for a brief description of key events 1–3. The Jensen Shannon Distance (JSD) is given in panel E and ranks the sampling strategies based on how similar the BDSKY estimates under those strategies are to those derived from *EpiFilter* (smaller values are better). Panel F provides the pairwise JSD between the BDSKY estimates under different sampling strategies, showing often appreciable difference among strategies.

#### Hong Kong

We applied the BDSKY model to estimate the *R*_*t*_ for each dataset subsampled according to the different sampling strategies (Figure 4). We compared these against the *R*_*t*_ from incidence data, derived from *EpiFilter*. Based on the proportional sampling scheme, which had the lowest JSD (Figure 4E), we initially infer a super-critical *R*_*t*_ value, with a mean around 2, that appears to fall swiftly in response to the state of emergency and the rapid implementation of NPIs. A steady transmission rate subsequently persisted throughout the following weeks around the critical threshold (*R*_*t*_ = 1). This period is followed by a sharp increase in *R*_*t*,_ peaking at a mean value of 2.6. This is likely due to imported cases from North America and Europe^68^. This led to a ban on international travel resulting in a sharp decline in *R*_*t*_ (Figure 2). However, this decline lasted around a week with the mean *R*_*t*_ briefly increasing until more stringent NPIs such as the banning of major gatherings were implemented. Following this, the *R*_*t*_ continued its sharp decline falling below the critical threshold, with transmission becoming sub-critical (Figure 4). The proportional sampling scheme showed the most divergence from all other sampling schemes whilst the uniform and reciprocal-proportional sampling schemes were almost identical (Figure 4F).

These results were mirrored in the estimation of *r*_*t*._ (Figure 5), where estimates derived from the proportional sampling scheme showed the least divergence (Figure 5E). There was an initial decline in the *r*_*t*_, which steadied at a value of ∼ 0, indicating that epidemic stabilisation had occurred. This stable period is followed by an increase in *r*_*t*_ peaking at around a 0.050 d^-1^ (Figure 5B). In response to NPIs, the *r*_*t*_ starts to decrease, falling below 0, indicating a receding epidemic. The rate of this decline peaks at around -0.075 d^-1^ (Figure 5B). Unlike the estimation of *R*_*t*_ (Figure 4), the unsampled sampling scheme showed the most divergence from all other sampling schemes (Figure 5F). It also has a high divergence from estimates derived from *EpiFilter* when compared the proportional sampling scheme which was the most closely related to *EpiFilter* (Figure 5E). Once again, the uniform and reciprocal-proportional schemes are the most closely related (Figure 5E).

**Figure 5:**
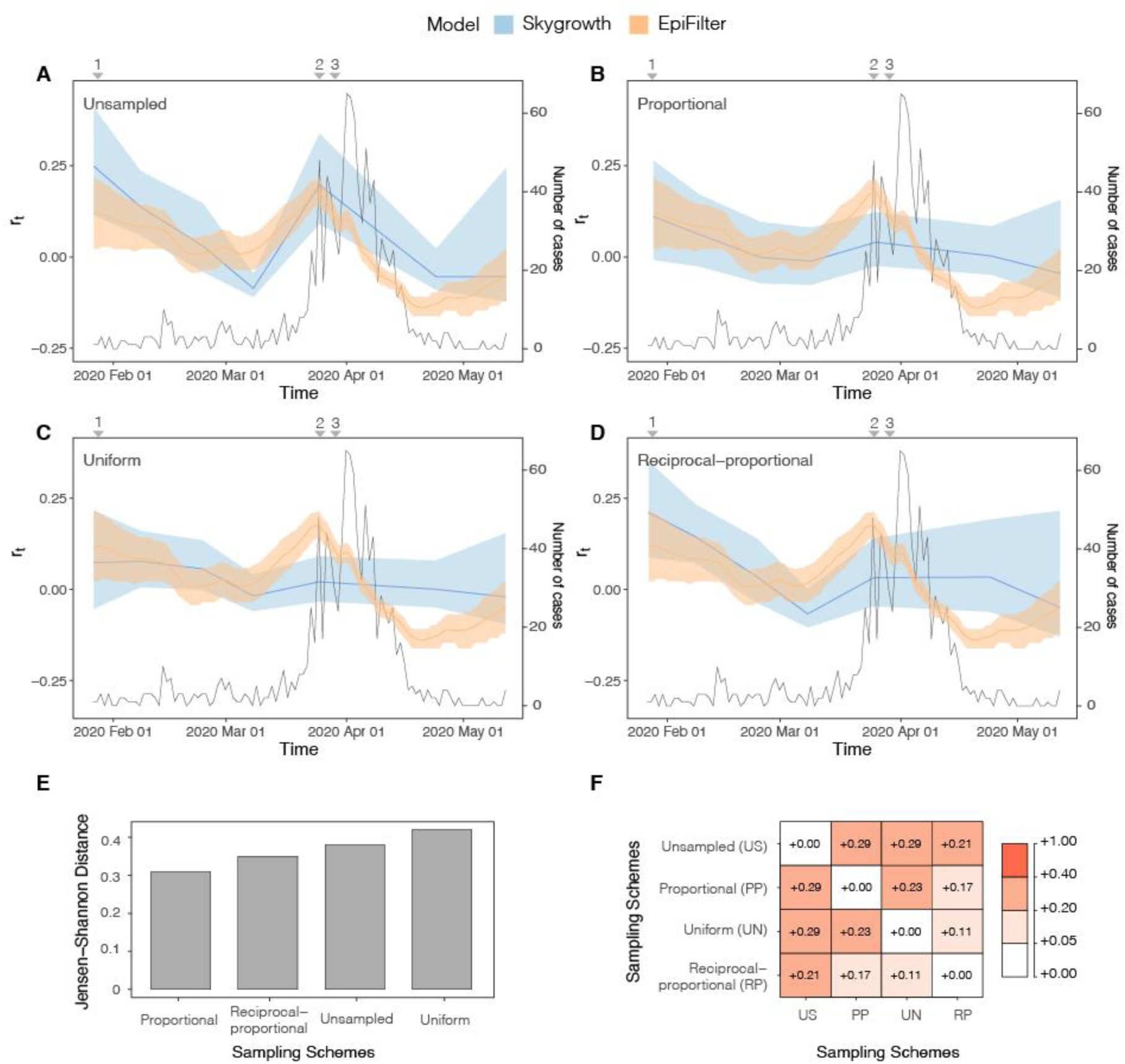
*r*_*t*_ estimated from both the *Skygrowth* and *EpiFilter* methods for Hong Kong. Titles indicate the sampling scheme used in panels A-D. The light-shaded area represents the 95% highest posterior density interval. The solid line represents the mean *r*_*t*_ estimate with *EpiFilter* in orange and *Skygrowth* in blue. The black line refers to the number of cases. We refer to Figure 1 for a brief description of key events 1–3. The Jensen Shannon Distance (JSD) is given in panel E and ranks the sampling strategies based on how similar the *Skygrowth* estimates under those strategies are to those derived from *EpiFilter* (smaller values are better). Panel F provides the pairwise JSD between the BDSKY estimates under different sampling strategies, showing often appreciable difference among strategies.

#### Brazil

The uniform, reciprocal-proportional, and proportional sampling schemes all showed a similarly low JSD (Figure 6E). Based on these sampling schemes, we initially infer super-critical transmission (*R*_*t*_ > 1) with a mean value of 3 (Figure 6). From this point, the *R*_*t*_ declines, although it remains above the critical threshold (*R*_*t*_ = 1) for much of the study period. Sub-critical transmission (*R*_*t*_ < 1) was only reached after the re-imposition of NPIs. This implies that initial restrictions, such as the suspension of commercial activities, were likely insufficient for suppressing spread. Only after more stringent restrictions were imposed did *R*_*t*_ become sub-critical. However, there is no evidence of a sharp decrease in *R*_*t*_ once restrictions were re-imposed, which may suggest limited effectiveness. The unsampled sampling scheme again showed the most divergence from all other sampling schemes (Figure 6F) and the highest divergence from the case data estimate (Figure 6E) with the uniform and proportional sampling schemes showing the most similarity. As such, applying no sampling strategy/opportunistic sampling leads to, from the perspective of comparing to *EpiFilter*, the most biased estimates.

**Figure 6:**
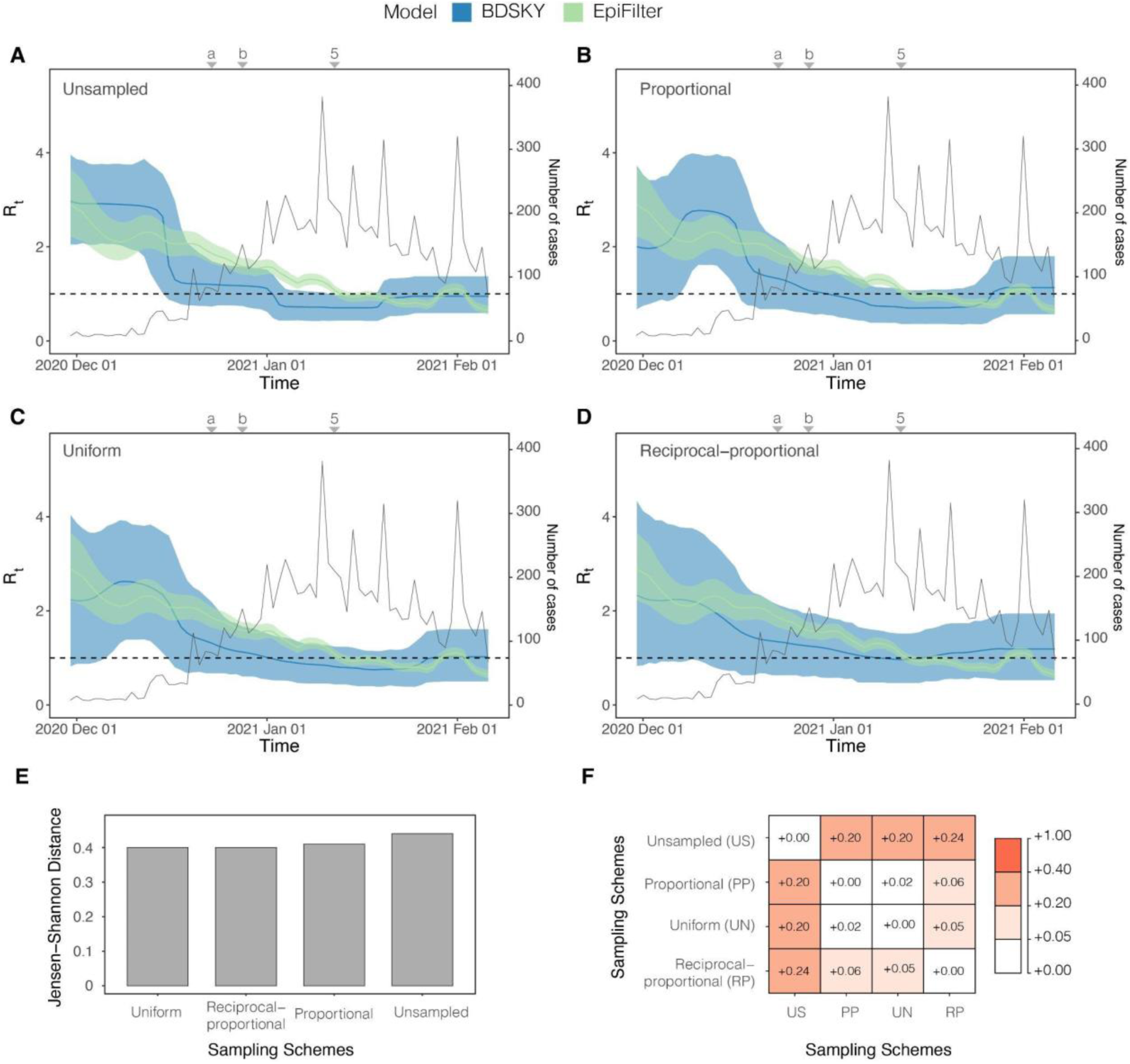
*R*_*t*_ estimated from both the BDSKY and *EpiFilter* methods forAmazonas, Brazil. Titles indicate the sampling scheme used in panels A-D. The light-shaded area represents the 95% highest posterior density interval. The solid line represents the mean *R*_*t*_ estimate with *EpiFilter* in green and BDSKY in blue. We refer to Figure 2 for a brief description of key events, including 5 which corresponds to the second lockdown. Event “a’’ corresponds to the suspension of commercial activities in Manaus; event “b” corresponds to the resumption of commercial activities in Manaus^72^. The Jensen Shannon Distance (JSD) is given in panel E and ranks the sampling strategies based on how similar the BDSKY estimates under those strategies are to those derived from *EpiFilter* (smaller values are better). Panel F provides the pairwise JSD between the BDSKY estimates under different sampling strategies, showing often appreciable difference among strategies.

Based on the proportional sampling scheme, which had the lowest JSD (Figure 7E) we infer a steady decline in *r*_*t*_ which matches the pattern seen with the *R*_*t*_ value (Figure 7). The initial *r*_*t*_ implied a 0.250 d^-1^. Subsequently, the *r*_*t*_ falls over the study period. *r*_*t*_ falls below 0 after the re-imposition of NPIs declining at -0.030 d^-1^ by the end of the study period. There is no evidence of any noticeable declines in *r*_*t*_ when interventions were introduced indicating that they might not have significantly impacted the growth rate of P.1/gamma. The unsampled sampling scheme was again most divergent from other sampling schemes as well as from estimates derived from *EpiFilter* with the uniform and reciprocal-proportional being most similar.

**Figure 7:**
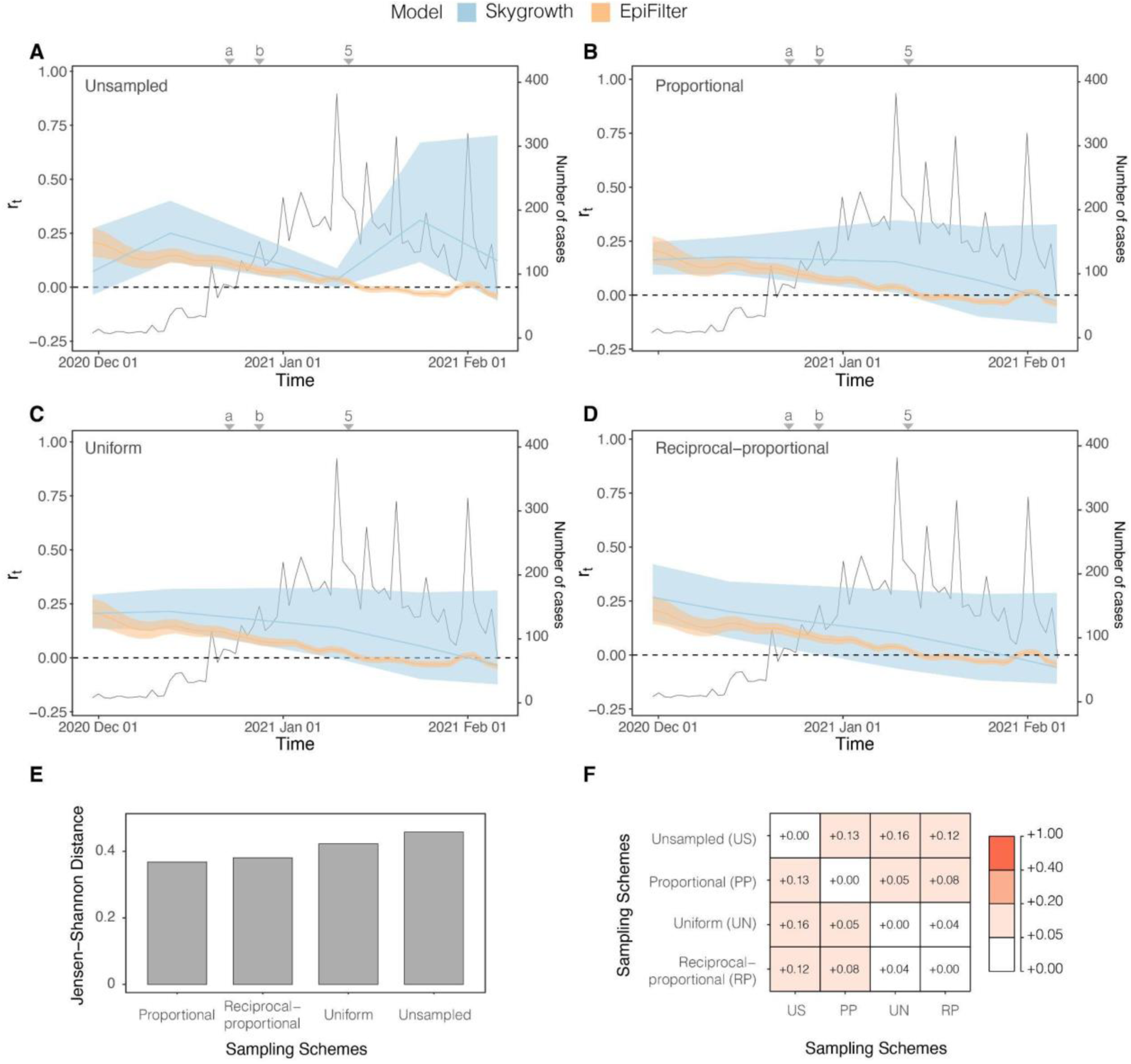
*r*_*t*_ estimated from both the *Skygrowth* and *EpiFilter* methods for Amazonas, Brazil. Titles indicate the sampling scheme used in panels A-D. The light-shaded area represents the 95% highest posterior density interval. The solid line represents the mean *r*_*t*_ estimate with *EpiFilter* in orange and *Skygrowth* in blue. We refer to Figure 2 for a brief description of key events, including 5 which corresponds to the second lockdown. Event “a’’ corresponds to the suspension of commercial activities in Manaus; event “b” corresponds to the resumption of commercial activities in Manaus^72^. The Jensen Shannon Distance (JSD) is given in panel E and ranks the sampling strategies based on how similar the *Skygrowth* estimates under those strategies are to those derived from *EpiFilter* (smaller values are better). Panel F provides the pairwise JSD between the BDSKY estimates under different sampling strategies, showing often appreciable difference among strategies.

## Discussion

In this study, we applied phylodynamic methods to available SARS-CoV-2 sequences from Hong Kong and the Amazonas state of Brazil to infer their key epidemiological parameters and to compare the impact that various sampling strategies have on the phylodynamic reconstruction of these parameters.

We estimated the basic reproductive number of SARS-CoV-2 in Hong Kong to be 2.17 (95% CI = 1.43-2.83). This supports previous estimates of the initial *R*_*0*_ in Hong Kong^68,74^ which estimated *R*_*0*_ to be 2.23 (95% CI = 1.47-3.42). For the Amazonas state in Brazil, we estimated the *R*_*0*_ to be 3.67 (95% CI = 2.83 – 4.48). Whilst the population of Amazonas State may not be fully susceptible to P.1/Gamma^14,82^, this should not affect the comparison among sampling schemes. We found that *R*_*0*_ is robust to changes in sampling schemes (Figure 3A and C).

For the Hong Kong dataset, the proportional sampling scheme was superior to all other sampling schemes in estimating *R*_*t*_. It successfully predicted the initial super-critical *R*_*t*_, its decline in response to rapid NPIs, and subsequent increase and decline during the second wave of infections (Figure 4B). This was in comparison to the uniform sampling scheme, which provided the worst (largest) JSD (Figure 4D) and an *R*_*t*_ estimate that was largely insensitive to NPIs. The proportional sampling scheme, alongside the uniform sampling scheme, best estimated *r*_*t*_ (Figure 5B and C). In contrast, for the Amazonas dataset, the uniform sampling scheme best estimated the *R*_*t*_ and *r*_*t*_ (Figure 6C) whilst the proportional sampling scheme best captured *r*_*t*_ (Figure 7C). It captured both its initial super-critical *R*_*t*_ and high *r*_*t*_ alongside their subsequent decline.

We found that estimates from all sampling schemes were distinct from those obtained using the unsampled data and that on some instances the sampling schemes were also appreciably different from one another (see panel F in Figures 4-7) with the uniform and reciprocal-proportional sampling strategies being most similar. This highlights how different sampling schemes can produce significantly differing estimates of epidemiological parameters and underscores the need for considering sampling and its potential impact on estimations.

Our *R*_*t*_ estimates are consistent with previous estimates of Gamma VOC’s transmissibility in Amazonas state^14^. This contrasted with the unsampled data in which the *r*_*t*_ increased at the end of the period (Figure 7A). This highlights that unlike *R*_*0*_, both *R*_*t*_ and *r*_*t*_ are sensitive to changes in sampling and that even related epidemiological parameters like *R*_*t*_ and *r*_*t*_ may require different sampling strategies to optimise inferences.

Molecular clock dating of the Hong Kong and Amazonas dataset has revealed that the date of origin is relatively robust to changes in sampling schemes. For Hong Kong, SARS-CoV-2 likely emerged in mid-December 2019 around 5 weeks before the first reported case on the 22^nd^ of January 2020^68^. The Amazonas dataset revealed that the date of the common ancestor of the P.1 lineage emerged around late October 2020 to early November, around 5 weeks before the first reported case on the 6^th^ of December^14^, with all BCI’s overlapping for each sampling strategy. Like the molecular clock dating, we found that the molecular clock rate was robust to changes in sampling strategies in both datasets with all sampling strategies having overlapped BCI’s (Supplementary Table 2 and Supplementary Figure 5). For the Hong Kong dataset, its clock rate is comparable to early estimations of the mean substitution rate per site per year of SARS-CoV-2^13^. However, the clock rate estimated for the Brazilian dataset is lower than the initial 8.00×10^−4^ s/s/y which is used in investigating SARS-CoV-2^75^ and that has been used in previous analyses of Gamma VOC^76^. This initial estimation of evolutionary rate was estimated from genomic data taken over a short time span at the beginning of the pandemic introducing a time dependency bias. By using a more appropriate clock rate it can improve tree height and rooting resulting in more robust parameter estimations^77^.

Treating sampling times as uninformative has been shown to be inferior to including them as dependent on effective population size and other parameters by several previous studies^30,31,34,78^. Whilst these studies did not consider the estimation of epidemiological parameters, they highlight the potential of systematic biases being introduced into the phylodynamic reconstruction by not using a sampling scheme or by assuming an incorrect model for how sampling schemes introduce information. This was supported by our results as phylodynamic inferences with no sampling strategy applied had the poorest overall performance for both Hong Kong and the Amazonas state. This implies that sampling design choices can significantly impact phylodynamic reconstruction, and that exploration of sampling strategies is needed to obtain the most reliable estimates of key epidemiological parameters.

While our results provide rigorous insight into the dynamics of SARS-CoV-2 and the impact of sampling strategies in the Amazonas state and Hong Kong, there are limitations. The *Skygrowth* and BDSKY models do not explicitly consider imports into their respective regions. This is particularly relevant for Hong Kong as most initial sequences from the region were sequenced from importation events^79^ which can introduce error into parameter estimation^80^. However, as the epidemic expanded, more infections were attributable to autochthonous transmission^79^, and the risk of error introduced by importation events decreased. Moreover, while sampling strategies can account for temporal variations in genomic sampling fractions there is currently no way to account for non-random sampling approaches in either the BDSKY or *Skygrowth* models^81^. It is unclear how network-based sampling may affect parameter estimates obtained through these models^82^ presenting a key challenge in molecular and genetic epidemiology. Spatial heterogeneities were also not explored within this work. This represents the next key step in understanding the impact of sampling as spatial sampling schemes would allow the reconstruction of the dispersal dynamics and estimation of epidemic overdispersion (*k*), a key epidemiological parameter.

Finally, we compared our phylodynamic estimates against epidemiological inferences derived from incident case data from Hong Kong and Amazonas state, two settings with very different diagnostic capacity. While Hong Kong has high quality case data with a high testing rate^69^, there is a large underreporting of SARS-CoV-2 cases in the Amazonas state^72,83^. Future epidemiological modelling work is needed to compare parameter estimates obtained from case data, death data and excess death data across different settings. This will improve the benchmarks we use to compare sequence-based estimates against.

This work has highlighted the impact and importance that applying temporal sampling strategies can have on phylodynamic reconstruction. Whilst more genomic datasets from a variety of countries and regions with different sampling intensities and proportions are needed to create a more generalisable sampling framework and to dissect any potential cofounders, this study has demonstrated that genomic datasets that commonly feature opportunistic sampling (i.e., there is no deliberate strategy design) can introduce significant uncertainty and biases in the estimation of epidemiological parameters. This finding signifies the need for more targeted attempts at performing genomic surveillance and epidemic analyses particularly in resource-poor settings with limited genomic capability.

## Data Availability

All data produced in the present study are available upon reasonable request to the authors

## CRediT authorship contribution statement

R.P.D.I, K.V.P and N.R.F conceived and designed the study, R.P.D.I wrote and performed the analyses. R.P.D.I wrote the manuscript which was edited and supervised by K.V.P and N.R.F. All authors have contributed to and approved the manuscript for submission.

## Supplementary Figures and Tables

**Supplementary Figure 1:**
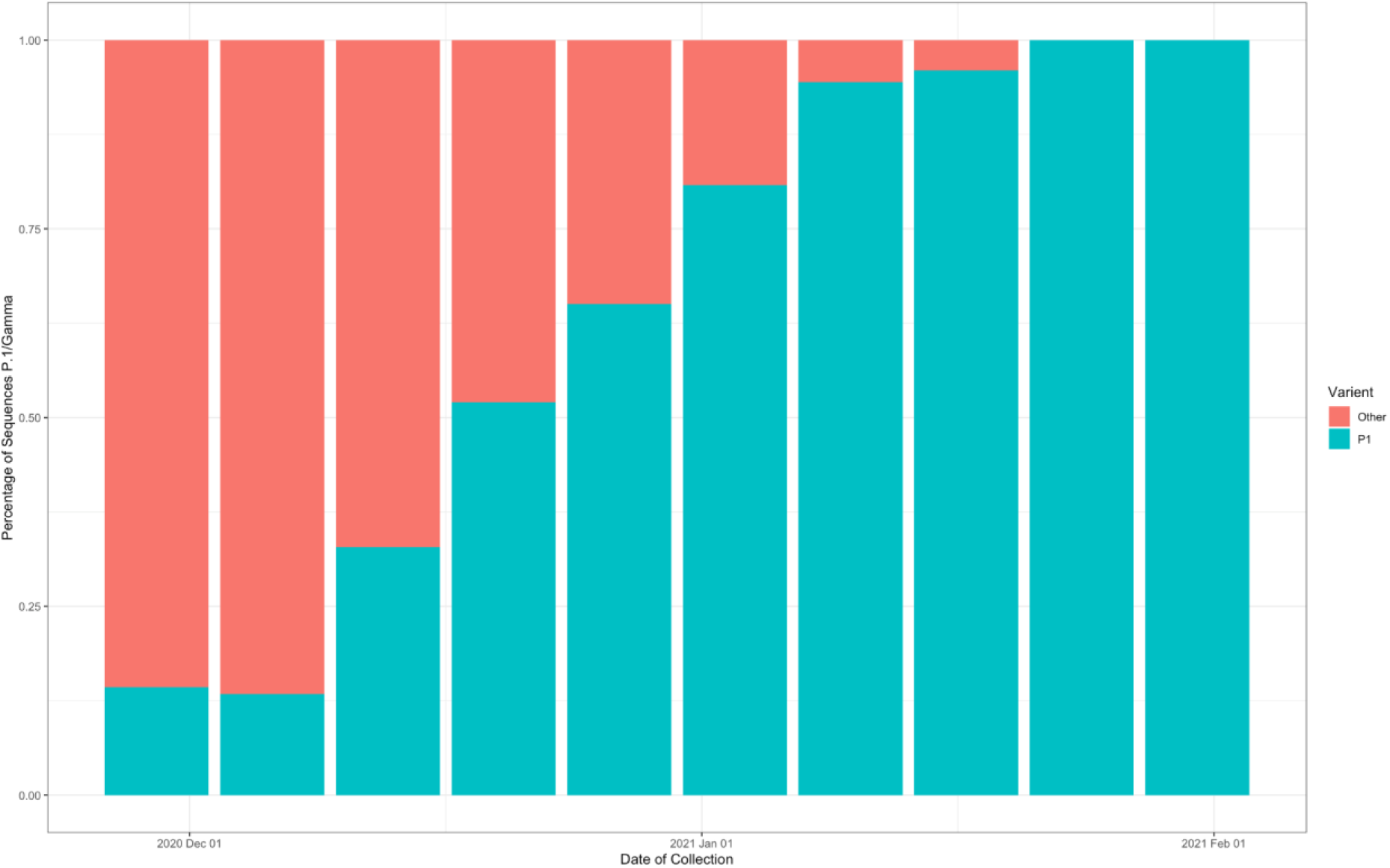
The proportion of P.1 sequences compared to non-P.1 sequences from Amazonas State, Brazil found on GISaid (Shu and McCauley, 2017).

**Supplementary Figure 2:**
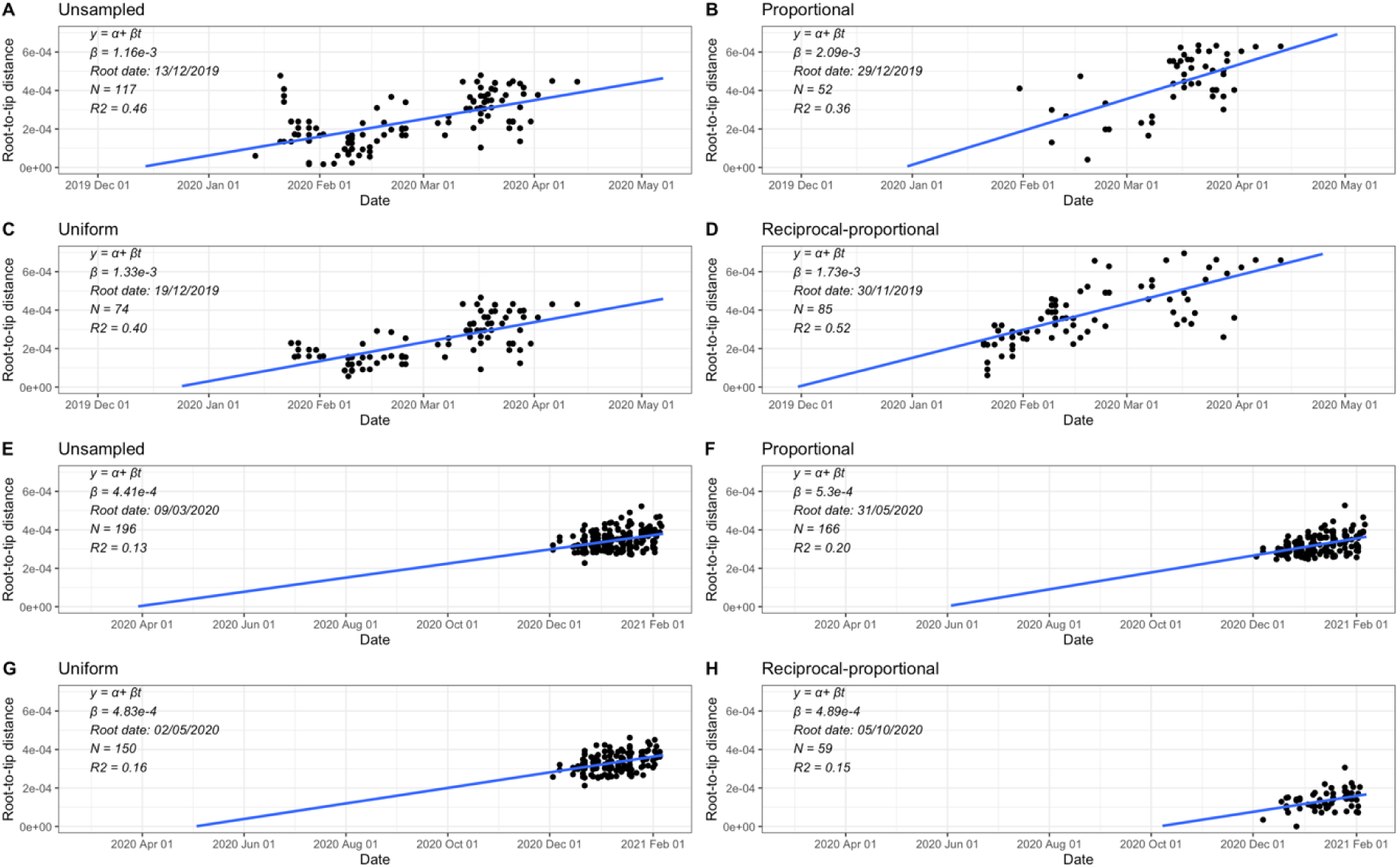
Root-to-tip genetic distances to sample collection dates for the SARS-CoV-2 genome datasets used in this study: A-D represents Hong Kong and E-H represent Amazonas State. Plots are based on the maximum likelihood trees rooted by maximising R^2^. The linear regression trend lines are shown to data points, corresponding to the genome sequences (represented with black dots).

**Supplementary Figure 3:**
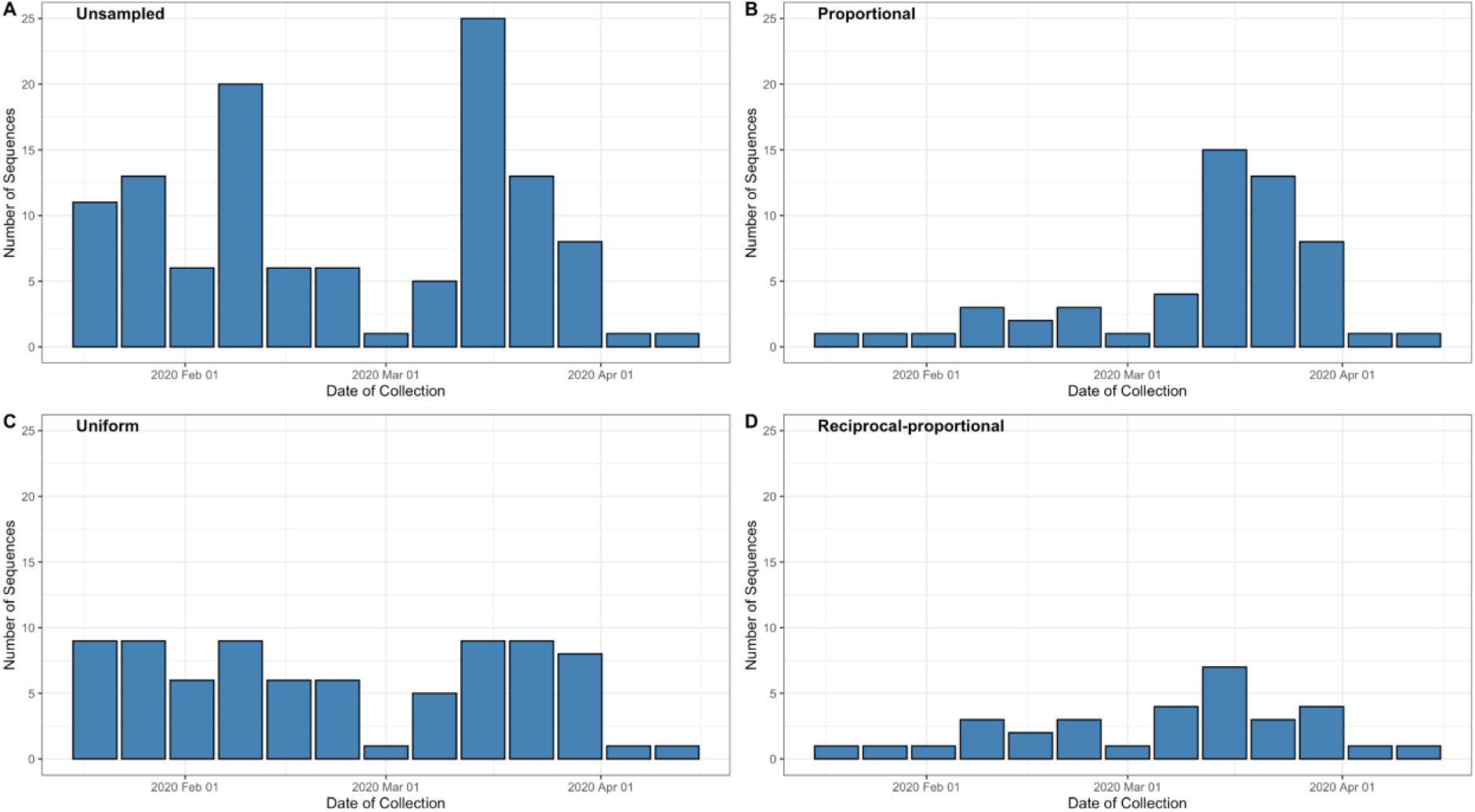
Number of sequences for each week and sampling scheme for Hong Kong dataset.

**Supplementary Figure 4:**
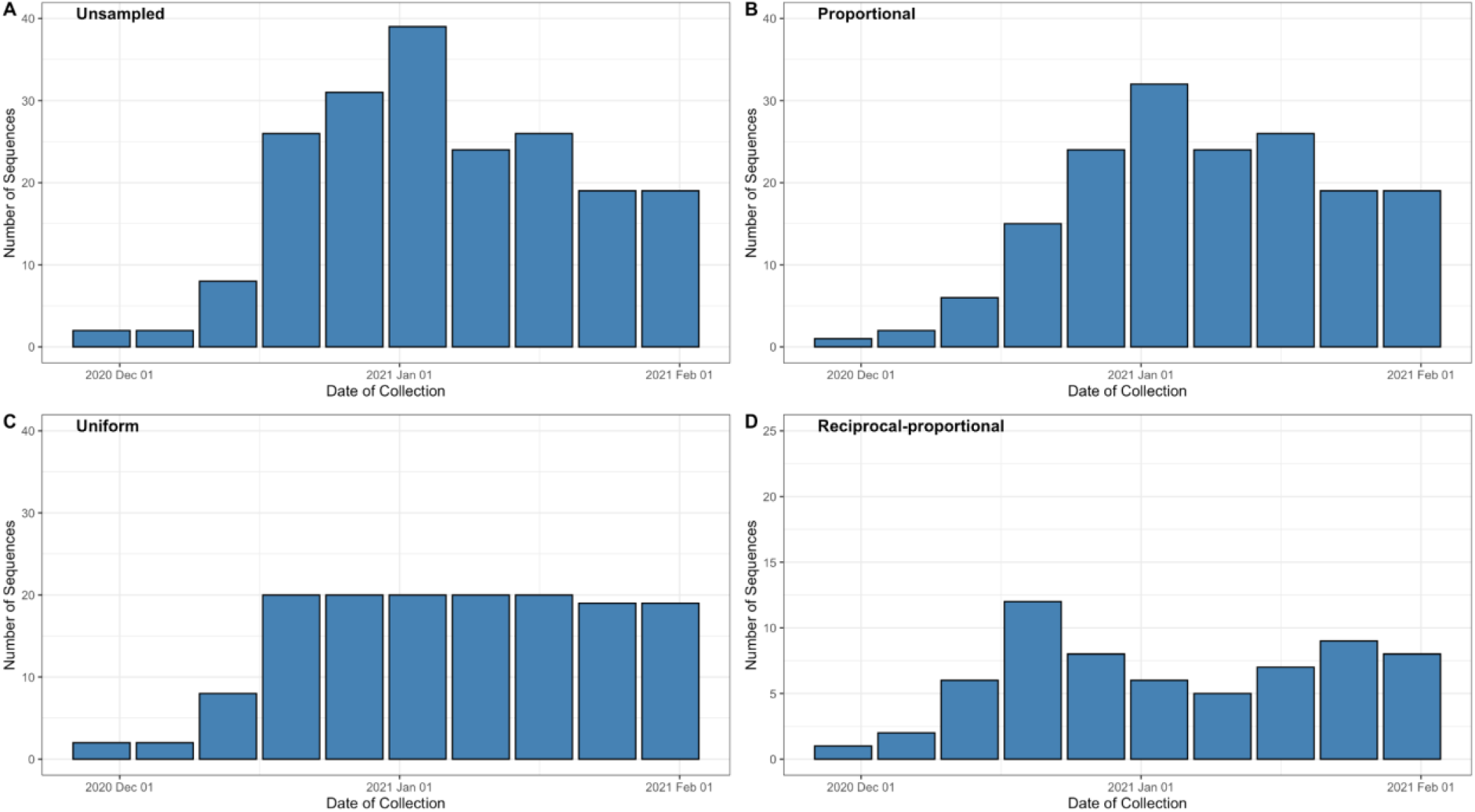
Number of sequences for each week and sampling scheme for Amazonas dataset.

**Supplementary Table 1:** TMRCA and mean substitution rate both with 95% BCI for each sampling strategy for Hong Kong and Amazonas datasets alongside the Jensen-Shannon distance. Full posterior distribution of the TMRCA and substitution rates obtained under the different sampling strategies can be found in Figure 3B and D and Supplementary Figure 5.

| **Sampling Strategy** | **Dataset** | **TMRCA (95% BCI)** | **Mean Substitution Rate (95% BCI, subs/site/year, s/s/y)** |
| --- | --- | --- | --- |
| Unsampled | Hong Kong | 2<sup>nd</sup> December 2019 (10<sup>th</sup> November 2019 – 24<sup>th</sup> December 2019) | 1.12x10<sup>-3</sup><br>(9.16x10<sup>-4</sup> – 1.35x10<sup>-3</sup>) |
|  | Brazil | 30<sup>th</sup> October 2020 (8<sup>th</sup> October 2020 – 13<sup>th</sup> December 2020) | 4.58x10<sup>-4</sup><br>(3.69x10<sup>-4</sup> – 5.56x10<sup>-4</sup>) |
| Proportional | Hong Kong | 24<sup>th</sup> December 2019 (21<sup>st</sup> November 2019 – 11<sup>th</sup> January 2020) | 1.39x10<sup>-3</sup><br>(9.28x10<sup>-4</sup> – 2.48x10<sup>-3</sup>) |
|  | Brazil | 30<sup>th</sup> October 2020 (25<sup>th</sup> August 2020 – 29<sup>th</sup> November 2020) | 4.60x10<sup>-4</sup><br>(3.70x10<sup>-4</sup> – 5.56x10<sup>-4</sup>) |
| Uniform | Hong Kong | 13<sup>th</sup> December 2019 (18<sup>th</sup> November 2019 – 4<sup>th</sup> January 2020) | 1.64x10<sup>-3</sup><br>(1.22x10<sup>-3</sup> – 2.09x10<sup>-3</sup>) |
| | Brazil | 27<sup>th</sup> October 2020 (5<sup>th</sup> October 2020 – 25<sup>th</sup> November 2020) | $4.60 \times 10^{-4}$<br>($3.70 \times 10^{-4}$ – $5.56 \times 10^{-4}$) |
| Reciprocal-proportional | Hong Kong | 6<sup>th</sup> December 2019 (10<sup>th</sup> November 2019 – 28<sup>th</sup> December 2019) | $1.30 \times 10^{-3}$<br>($1.03 \times 10^{-3}$ – $1.59 \times 10^{-3}$) |
| | Brazil | 30<sup>th</sup> October 2020 (27<sup>th</sup> September 2020 – 25<sup>th</sup> November 2020) | $4.00 \times 10^{-4}$<br>($2.56 \times 10^{-4}$ – $5.55 \times 10^{-4}$) |

**Supplementary Figure 5:**
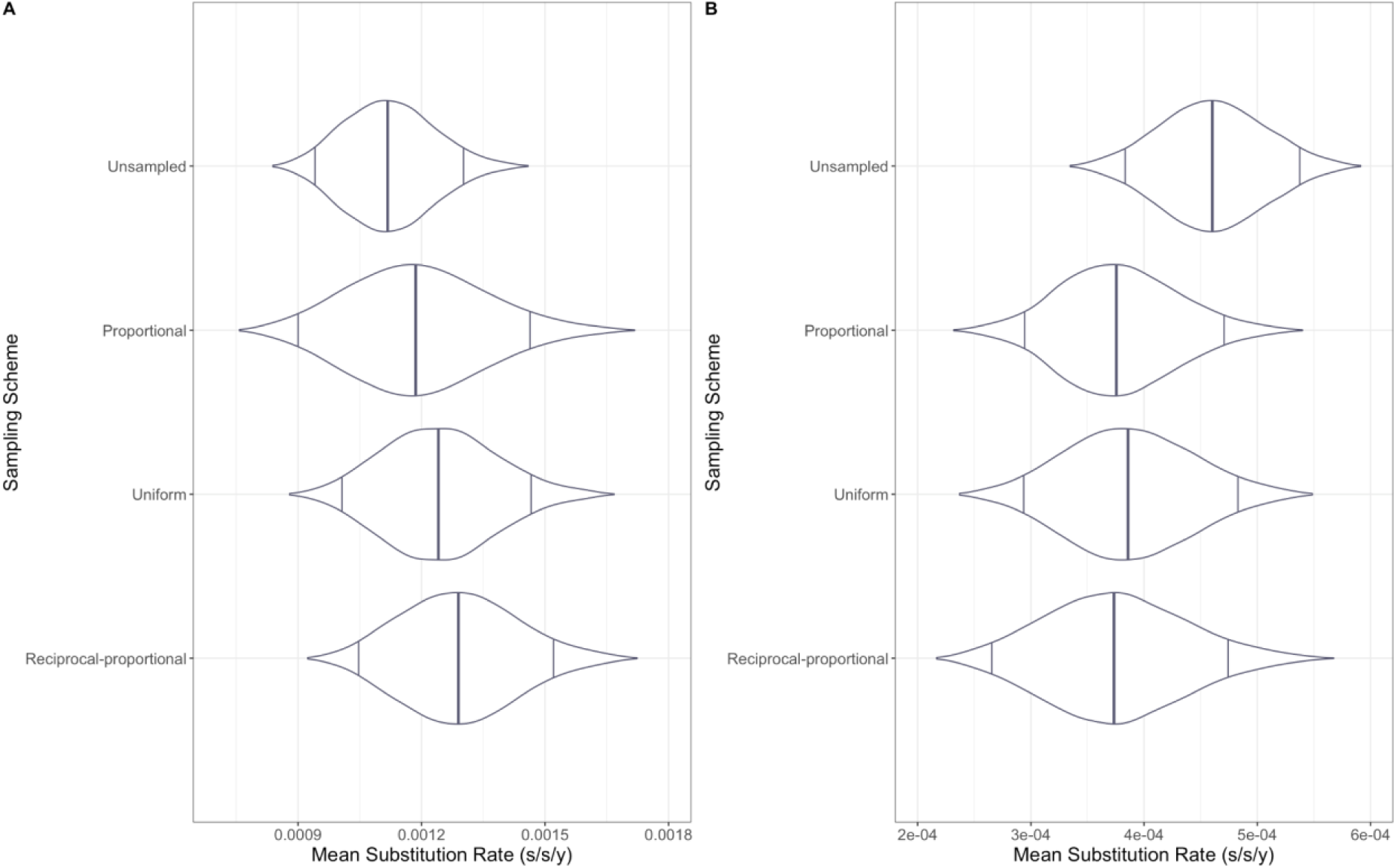
Mean substitution rate (s/s/y) for Hong Kong and Brazil. Figure 1A represents Hong Kong with Figure 1B representing the Amazonas. The central line represents the posterior mean and with intervals representing 95% Highest Posterior Density Interval

**Supplementary Table 2:** Accession ID of each Hong Kong sequence for each sampling strategy used within this study

| Unsampled | Proportional | Uniform | Reciprocal-proportional |
| --- | --- | --- | --- |
| EPI_ISL_ 412028 | EPI_ISL_ 414517 | EPI_ISL_ 412029 | EPI_ISL_ 412028 |
| EPI_ISL_ 412029 | EPI_ISL_ 414519 | EPI_ISL_ 414517 | EPI_ISL_ 412029 |
| EPI_ISL_ 412030 | EPI_ISL_ 414527 | EPI_ISL_ 414519 | EPI_ISL_ 412030 |
| EPI_ISL_ 414517 | EPI_ISL_ 418815 | EPI_ISL_ 414527 | EPI_ISL_ 414517 |
| EPI_ISL_ 414519 | EPI_ISL_ 419224 | EPI_ISL_ 414569 | EPI_ISL_ 414519 |
| EPI_ISL_ 414527 | EPI_ISL_ 419229 | EPI_ISL_ 414571 | EPI_ISL_ 414527 |
| EPI_ISL_ 414528 | EPI_ISL_ 419232 | EPI_ISL_ 416314 | EPI_ISL_ 414528 |
| EPI_ISL_ 414569 | EPI_ISL_ 450404 | EPI_ISL_ 417064 | EPI_ISL_ 414569 |
| EPI_ISL_ 414571 | EPI_ISL_ 450405 | EPI_ISL_ 417443 | EPI_ISL_ 414571 |
| EPI_ISL_ 416314 | EPI_ISL_ 450410 | EPI_ISL_ 419214 | EPI_ISL_ 416314 |
| EPI_ISL_ 417064 | EPI_ISL_ 476801 | EPI_ISL_ 419215 | EPI_ISL_ 417064 |
| EPI_ISL_ 417176 | EPI_ISL_ 476802 | EPI_ISL_ 419217 | EPI_ISL_ 417176 |
| EPI_ISL_ 417178 | EPI_ISL_ 476803 | EPI_ISL_ 419224 | EPI_ISL_ 417178 |
| EPI_ISL_ 417181 | EPI_ISL_ 497769 | EPI_ISL_ 419225 | EPI_ISL_ 417181 |
| EPI_ISL_ 417185 | EPI_ISL_ 497773 | EPI_ISL_ 419227 | EPI_ISL_ 417185 |
| EPI_ISL_ 417187 | EPI_ISL_ 497775 | EPI_ISL_ 419228 | EPI_ISL_ 417187 |
| EPI_ISL_ 417188 | EPI_ISL_ 497784 | EPI_ISL_ 419229 | EPI_ISL_ 417188 |
| EPI_ISL_ 417193 | EPI_ISL_ 497786 | EPI_ISL_ 419231 | EPI_ISL_ 417193 |
| EPI_ISL_ 417197 | EPI_ISL_ 497791 | EPI_ISL_ 419232 | EPI_ISL_ 417197 |
| EPI_ISL_ 417443 | EPI_ISL_ 497796 | EPI_ISL_ 419245 | EPI_ISL_ 417443 |
| EPI_ISL_ 418815 | EPI_ISL_ 497799 | EPI_ISL_ 419247 | EPI_ISL_ 418815 |
| EPI_ISL_ 419214 | EPI_ISL_ 497806 | EPI_ISL_ 419250 | EPI_ISL_ 419214 |
| EPI_ISL_ 419215 | EPI_ISL_ 497808 | EPI_ISL_ 419252 | EPI_ISL_ 419215 |
| EPI_ISL_ 419216 | EPI_ISL_ 497810 | EPI_ISL_ 434564 | EPI_ISL_ 419216 |
| EPI_ISL_ 419217 | EPI_ISL_ 497811 | EPI_ISL_ 434565 | EPI_ISL_ 419217 |
| EPI_ISL_ 419219 | EPI_ISL_ 497818 | EPI_ISL_ 434567 | EPI_ISL_ 419219 |
| EPI_ISL_ 419221 | EPI_ISL_ 497819 | EPI_ISL_ 434568 | EPI_ISL_ 419221 |
| EPI_ISL_ 419222 | EPI_ISL_ 497821 | EPI_ISL_ 434569 | EPI_ISL_ 419222 |
| EPI_ISL_ 419224 | EPI_ISL_ 497823 | EPI_ISL_ 434570 | EPI_ISL_ 419224 |
| EPI_ISL_ 419225 | EPI_ISL_ 497824 | EPI_ISL_ 434571 | EPI_ISL_ 419225 |
| EPI_ISL_ 419226 | EPI_ISL_ 497840 | EPI_ISL_ 450405 | EPI_ISL_ 419226 |
| EPI_ISL_ 419227 | EPI_ISL_ 497845 | EPI_ISL_ 450408 | EPI_ISL_ 419227 |
| EPI_ISL_ 419228 | EPI_ISL_ 497846 | EPI_ISL_ 450409 | EPI_ISL_ 419228 |
| EPI_ISL_ 419229 | EPI_ISL_ 497847 | EPI_ISL_ 450410 | EPI_ISL_ 419229 |
| EPI_ISL_ 419231 | EPI_ISL_ 497850 | EPI_ISL_ 450411 | EPI_ISL_ 419231 |
| EPI_ISL_ 419232 | EPI_ISL_ 497856 | EPI_ISL_ 476801 | EPI_ISL_ 419232 |
| EPI_ISL_ 419245 | EPI_ISL_ 497865 | EPI_ISL_ 476802 | EPI_ISL_ 419245 |
| EPI_ISL_ 419247 | EPI_ISL_ 497870 | EPI_ISL_ 476804 | EPI_ISL_ 419247 |
| EPI_ISL_ 419250 | EPI_ISL_ 516798 | EPI_ISL_ 497769 | EPI_ISL_ 419250 |
| EPI_ISL_ 419252 | EPI_ISL_ 539820 | EPI_ISL_ 497771 | EPI_ISL_ 419252 |
| EPI_ISL_ 434560 | EPI_ISL_ 539850 | EPI_ISL_ 497783 | EPI_ISL_ 434563 |
| EPI_ISL_ 434563 | EPI_ISL_ 539851 | EPI_ISL_ 497784 | EPI_ISL_ 434564 |
| EPI_ISL_ 434564 | EPI_ISL_ 610167 | EPI_ISL_ 497791 | EPI_ISL_ 434565 |
| EPI_ISL_ 434565 | EPI_ISL_ 610168 | EPI_ISL_ 497806 | EPI_ISL_ 434566 |
| EPI_ISL_ 434566 | EPI_ISL_ 610169 | EPI_ISL_ 497810 | EPI_ISL_ 434567 |
| EPI_ISL_ 434567 | EPI_ISL_ 610170 | EPI_ISL_ 497811 | EPI_ISL_ 434568 |
| EPI_ISL_ 434568 | EPI_ISL_ 610171 | EPI_ISL_ 497813 | EPI_ISL_ 434569 |
| EPI_ISL_ 434569 | EPI_ISL_ 610172 | EPI_ISL_ 497818 | EPI_ISL_ 434570 |
| EPI_ISL_ 434570 | EPI_ISL_ 610173 | EPI_ISL_ 497821 | EPI_ISL_ 434571 |
| EPI_ISL_ 434571 | EPI_ISL_ 610174 | EPI_ISL_ 497823 | EPI_ISL_ 450405 |
| EPI_ISL_ 450404 | EPI_ISL_ 610175 | EPI_ISL_ 497824 | EPI_ISL_ 450408 |
| EPI_ISL_ 450405 | EPI_ISL_ 610177 | EPI_ISL_ 497826 | EPI_ISL_ 450409 |
| EPI_ISL_ 450408 |  | EPI_ISL_ 497827 | EPI_ISL_ 450410 |
| EPI_ISL_ 450409 |  | EPI_ISL_ 497831 | EPI_ISL_ 450411 |
| EPI_ISL_ 450410 |  | EPI_ISL_ 497832 | EPI_ISL_ 450412 |
| EPI_ISL_ 450411 |  | EPI_ISL_ 497846 | EPI_ISL_ 476802 |
| EPI_ISL_ 450412 |  | EPI_ISL_ 497847 | EPI_ISL_ 476804 |
| EPI_ISL_ 476801 |  | EPI_ISL_ 497848 | EPI_ISL_ 497769 |
| EPI_ISL_ 476802 |  | EPI_ISL_ 497856 | EPI_ISL_ 497771 |
| EPI_ISL_ 476803 |  | EPI_ISL_ 497860 | EPI_ISL_ 497773 |
| EPI_ISL_ 476804 |  | EPI_ISL_ 497865 | EPI_ISL_ 497783 |
| EPI_ISL_ 497769 |  | EPI_ISL_ 539820 | EPI_ISL_ 497784 |
| EPI_ISL_ 497771 |  | EPI_ISL_ 539850 | EPI_ISL_ 497791 |
| EPI_ISL_ 497773 |  | EPI_ISL_ 539851 | EPI_ISL_ 497797 |
| EPI_ISL_ 497775 |  | EPI_ISL_ 610165 | EPI_ISL_ 497811 |
| EPI_ISL_ 497783 |  | EPI_ISL_ 610166 | EPI_ISL_ 497812 |
| EPI_ISL_ 497784 |  | EPI_ISL_ 610167 | EPI_ISL_ 497818 |
| EPI_ISL_ 497786 |  | EPI_ISL_ 610168 | EPI_ISL_ 497819 |
| EPI_ISL_ 497791 |  | EPI_ISL_ 610169 | EPI_ISL_ 497823 |
| EPI_ISL_ 497796 |  | EPI_ISL_ 610171 | EPI_ISL_ 497824 |
| EPI_ISL_ 497797 |  | EPI_ISL_ 610173 | EPI_ISL_ 497827 |
| EPI_ISL_ 497798 |  | EPI_ISL_ 610174 | EPI_ISL_ 497831 |
| EPI_ISL_ 497799 |  | EPI_ISL_ 610175 | EPI_ISL_ 497833 |
| EPI_ISL_ 497806 |  | EPI_ISL_ 610177 | EPI_ISL_ 497848 |
| EPI_ISL_ 497808 |  |  | EPI_ISL_ 497850 |
| EPI_ISL_ 497810 |  |  | EPI_ISL_ 497856 |
| EPI_ISL_ 497811 |  |  | EPI_ISL_ 497860 |
| EPI_ISL_ 497812 |  |  | EPI_ISL_ 497864 |
| EPI_ISL_ 497813 |  |  | EPI_ISL_ 497865 |
| EPI_ISL_ 497818 |  |  | EPI_ISL_ 539850 |
| EPI_ISL_ 497819 |  |  | EPI_ISL_ 539851 |
| EPI_ISL_ 497820 |  |  | EPI_ISL_ 610165 |
| EPI_ISL_ 497821 |  |  | EPI_ISL_ 610166 |
| EPI_ISL_ 497823 |  |  | EPI_ISL_ 610172 |
| EPI_ISL_ 497824 |  |  | EPI_ISL_ 610177 |
| EPI_ISL_ 497826 |  |  |  |
| EPI_ISL_ 497827 |  |  |  |
| EPI_ISL_ 497831 |  |  |  |
| EPI_ISL_ 497832 |  |  |  |
| EPI_ISL_ 497833 |  |  |  |
| EPI_ISL_ 497840 |  |  |  |
| EPI_ISL_ 497845 |  |  |  |
| EPI_ISL_ 497846 |  |  |  |
| EPI_ISL_ 497847 |  |  |  |
| EPI_ISL_ 497848 |  |  |  |
| EPI_ISL_ 497850 |  |  |  |
| EPI_ISL_ 497856 |  |  |  |
| EPI_ISL_ 497860 |  |  |  |
| EPI_ISL_ 497864 |  |  |  |
| EPI_ISL_ 497865 |  |  |  |
| EPI_ISL_ 497870 |  |  |  |
| EPI_ISL_ 516798 |  |  |  |
| EPI_ISL_ 539820 |  |  |  |
| EPI_ISL_ 539850 |  |  |  |
| EPI_ISL_ 539851 |  |  |  |
| EPI_ISL_ 610165 |  |  |  |
| EPI_ISL_ 610166 |  |  |  |
| EPI_ISL_ 610167 |  |  |  |
| EPI_ISL_ 610168 |  |  |  |
| EPI_ISL_ 610169 |  |  |  |
| EPI_ISL_ 610170 |  |  |  |
| EPI_ISL_ 610171 |  |  |  |
| EPI_ISL_ 610172 |  |  |  |
| EPI_ISL_ 610173 |  |  |  |
| EPI_ISL_ 610174 |  |  |  |
| EPI_ISL_ 610175 |  |  |  |
| EPI_ISL_ 610177 |  |  |  |

**Supplementary Table 3:** Accession ID of each Amazonas State, Brazil sequence for each sampling strategy used within this study

| Unsampled | Proportional | Uniform | Reciprocal-proportional |
| --- | --- | --- | --- |
| EPI_ISL_ 1034306 | EPI_ISL_ 1034304 | EPI_ISL_ 1034304 | EPI_ISL_ 1034306 |
| EPI_ISL_ 1060876 | EPI_ISL_ 1034306 | EPI_ISL_ 1034306 | EPI_ISL_ 1060913 |
| EPI_ISL_ 1060877 | EPI_ISL_ 1060877 | EPI_ISL_ 1060877 | EPI_ISL_ 1060914 |
| EPI_ISL_ 1060881 | EPI_ISL_ 1060881 | EPI_ISL_ 1060881 | EPI_ISL_ 1068149 |
| EPI_ISL_ 1060888 | EPI_ISL_ 1060897 | EPI_ISL_ 1060888 | EPI_ISL_ 1068150 |
| EPI_ISL_ 1060889 | EPI_ISL_ 1060900 | EPI_ISL_ 1060889 | EPI_ISL_ 1068156 |
| EPI_ISL_ 1060894 | EPI_ISL_ 1060902 | EPI_ISL_ 1060897 | EPI_ISL_ 1068198 |
| EPI_ISL_ 1060897 | EPI_ISL_ 1060904 | EPI_ISL_ 1060900 | EPI_ISL_ 1068258 |
| EPI_ISL_ 1060900 | EPI_ISL_ 1060906 | EPI_ISL_ 1060912 | EPI_ISL_ 1068260 |
| EPI_ISL_ 1060902 | EPI_ISL_ 1060912 | EPI_ISL_ 1060913 | EPI_ISL_ 1068262 |
| EPI_ISL_ 1060904 | EPI_ISL_ 1060913 | EPI_ISL_ 1060956 | EPI_ISL_ 1068263 |
| EPI_ISL_ 1060906 | EPI_ISL_ 1060914 | EPI_ISL_ 1061026 | EPI_ISL_ 1068264 |
| EPI_ISL_ 1060911 | EPI_ISL_ 1060918 | EPI_ISL_ 1068111 | EPI_ISL_ 1068278 |
| EPI_ISL_ 1060912 | EPI_ISL_ 1060956 | EPI_ISL_ 1068149 | EPI_ISL_ 1068286 |
| EPI_ISL_ 1060913 | EPI_ISL_ 1061026 | EPI_ISL_ 1068150 | EPI_ISL_ 1068288 |
| EPI_ISL_ 1060914 | EPI_ISL_ 1068110 | EPI_ISL_ 1068154 | EPI_ISL_ 1166615 |
| EPI_ISL_ 1060918 | EPI_ISL_ 1068111 | EPI_ISL_ 1068158 | EPI_ISL_ 1213190 |
| EPI_ISL_ 1060956 | EPI_ISL_ 1068112 | EPI_ISL_ 1068160 | EPI_ISL_ 1261690 |
| EPI_ISL_ 1061026 | EPI_ISL_ 1068114 | EPI_ISL_ 1068169 | EPI_ISL_ 1261694 |
| EPI_ISL_ 1068110 | EPI_ISL_ 1068149 | EPI_ISL_ 1068198 | EPI_ISL_ 2777236 |
| EPI_ISL_ 1068111 | EPI_ISL_ 1068150 | EPI_ISL_ 1068222 | EPI_ISL_ 2777320 |
| EPI_ISL_ 1068112 | EPI_ISL_ 1068151 | EPI_ISL_ 1068225 | EPI_ISL_ 2777363 |
| EPI_ISL_ 1068114 | EPI_ISL_ 1068154 | EPI_ISL_ 1068226 | EPI_ISL_ 2777375 |
| EPI_ISL_ 1068149 | EPI_ISL_ 1068156 | EPI_ISL_ 1068243 | EPI_ISL_ 2777376 |
| EPI_ISL_ 1068150 | EPI_ISL_ 1068158 | EPI_ISL_ 1068248 | EPI_ISL_ 2777384 |
| EPI_ISL_ 1068151 | EPI_ISL_ 1068160 | EPI_ISL_ 1068249 | EPI_ISL_ 2777388 |
| EPI_ISL_ 1068154 | EPI_ISL_ 1068169 | EPI_ISL_ 1068260 | EPI_ISL_ 2777397 |
| EPI_ISL_ 1068156 | EPI_ISL_ 1068198 | EPI_ISL_ 1068261 | EPI_ISL_ 2777399 |
| EPI_ISL_ 1068158 | EPI_ISL_ 1068221 | EPI_ISL_ 1068262 | EPI_ISL_ 2777401 |
| EPI_ISL_ 1068160 | EPI_ISL_ 1068222 | EPI_ISL_ 1068263 | EPI_ISL_ 2777403 |
| EPI_ISL_ 1068169 | EPI_ISL_ 1068225 | EPI_ISL_ 1068264 | EPI_ISL_ 2777404 |
| EPI_ISL_ 1068198 | EPI_ISL_ 1068248 | EPI_ISL_ 1068266 | EPI_ISL_ 2777409 |
| EPI_ISL_ 1068221 | EPI_ISL_ 1068249 | EPI_ISL_ 1068268 | EPI_ISL_ 2777410 |
| EPI_ISL_ 1068222 | EPI_ISL_ 1068258 | EPI_ISL_ 1068269 | EPI_ISL_ 2777414 |
| EPI_ISL_ 1068225 | EPI_ISL_ 1068260 | EPI_ISL_ 1068270 | EPI_ISL_ 2777415 |
| EPI_ISL_ 1068226 | EPI_ISL_ 1068261 | EPI_ISL_ 1068271 | EPI_ISL_ 2777465 |
| EPI_ISL_ 1068243 | EPI_ISL_ 1068262 | EPI_ISL_ 1068272 | EPI_ISL_ 2777466 |
| EPI_ISL_ 1068248 | EPI_ISL_ 1068263 | EPI_ISL_ 1068273 | EPI_ISL_ 2777467 |
| EPI_ISL_ 1068249 | EPI_ISL_ 1068264 | EPI_ISL_ 1068274 | EPI_ISL_ 2777469 |
| EPI_ISL_ 1068258 | EPI_ISL_ 1068266 | EPI_ISL_ 1068279 | EPI_ISL_ 2777470 |
| EPI_ISL_ 1068260 | EPI_ISL_ 1068268 | EPI_ISL_ 1068282 | EPI_ISL_ 2777472 |
| EPI_ISL_ 1068261 | EPI_ISL_ 1068269 | EPI_ISL_ 1068283 | EPI_ISL_ 2777473 |
| EPI_ISL_ 1068262 | EPI_ISL_ 1068270 | EPI_ISL_ 1068284 | EPI_ISL_ 2777474 |
| EPI_ISL_ 1068263 | EPI_ISL_ 1068271 | EPI_ISL_ 1068285 | EPI_ISL_ 2777475 |
| EPI_ISL_ 1068264 | EPI_ISL_ 1068272 | EPI_ISL_ 1068286 | EPI_ISL_ 2777482 |
| EPI_ISL_ 1068266 | EPI_ISL_ 1068273 | EPI_ISL_ 1068287 | EPI_ISL_ 2777483 |
| EPI_ISL_ 1068268 | EPI_ISL_ 1068274 | EPI_ISL_ 1068288 | EPI_ISL_ 2777485 |
| EPI_ISL_ 1068269 | EPI_ISL_ 1068275 | EPI_ISL_ 1068290 | EPI_ISL_ 2777503 |
| EPI_ISL_ 1068270 | EPI_ISL_ 1068276 | EPI_ISL_ 1068291 | EPI_ISL_ 2777508 |
| EPI_ISL_ 1068271 | EPI_ISL_ 1068278 | EPI_ISL_ 1068292 | EPI_ISL_ 2777509 |
| EPI_ISL_ 1068272 | EPI_ISL_ 1068279 | EPI_ISL_ 1166615 | EPI_ISL_ 2777516 |
| EPI_ISL_ 1068273 | EPI_ISL_ 1068280 | EPI_ISL_ 1213190 | EPI_ISL_ 2777599 |
| EPI_ISL_ 1068274 | EPI_ISL_ 1068281 | EPI_ISL_ 1213204 | EPI_ISL_ 2777698 |
| EPI_ISL_ 1068275 | EPI_ISL_ 1068282 | EPI_ISL_ 1261683 | EPI_ISL_ 2777986 |
| EPI_ISL_ 1068276 | EPI_ISL_ 1068283 | EPI_ISL_ 1261685 | EPI_ISL_ 2777987 |
| EPI_ISL_ 1068278 | EPI_ISL_ 1068284 | EPI_ISL_ 1261690 | EPI_ISL_ 2777993 |
| EPI_ISL_ 1068279 | EPI_ISL_ 1068285 | EPI_ISL_ 1261694 | EPI_ISL_ 2777999 |
| EPI_ISL_ 1068280 | EPI_ISL_ 1068286 | EPI_ISL_ 2777236 | EPI_ISL_ 2778002 |
| EPI_ISL_ 1068281 | EPI_ISL_ 1068287 | EPI_ISL_ 2777248 | EPI_ISL_ 2778004 |
| EPI_ISL_ 1068282 | EPI_ISL_ 1068288 | EPI_ISL_ 2777249 | EPI_ISL_ 2778005 |
| EPI_ISL_ 1068283 | EPI_ISL_ 1068289 | EPI_ISL_ 2777250 | EPI_ISL_ 833138 |
| EPI_ISL_ 1068284 | EPI_ISL_ 1068290 | EPI_ISL_ 2777320 | EPI_ISL_ 833140 |
| EPI_ISL_ 1068285 | EPI_ISL_ 1068291 | EPI_ISL_ 2777363 | EPI_ISL_ 906071 |
| EPI_ISL_ 1068286 | EPI_ISL_ 1068292 | EPI_ISL_ 2777364 | EPI_ISL_ 918505 |
| EPI_ISL_ 1068287 | EPI_ISL_ 1166615 | EPI_ISL_ 2777373 | EPI_ISL_ 918506 |
| EPI_ISL_ 1068288 | EPI_ISL_ 1213190 | EPI_ISL_ 2777374 | EPI_ISL_ 918508 |
| EPI_ISL_ 1068289 | EPI_ISL_ 1213204 | EPI_ISL_ 2777375 | EPI_ISL_ 918509 |
| EPI_ISL_ 1068290 | EPI_ISL_ 1261683 | EPI_ISL_ 2777376 |  |
| EPI_ISL_ 1068291 | EPI_ISL_ 1261685 | EPI_ISL_ 2777377 |  |
| EPI_ISL_ 1068292 | EPI_ISL_ 1261690 | EPI_ISL_ 2777378 |  |
| EPI_ISL_ 1166615 | EPI_ISL_ 1261694 | EPI_ISL_ 2777380 |  |
| EPI_ISL_ 1213190 | EPI_ISL_ 2777236 | EPI_ISL_ 2777383 |  |
| EPI_ISL_ 1213204 | EPI_ISL_ 2777238 | EPI_ISL_ 2777384 |  |
| EPI_ISL_ 1261683 | EPI_ISL_ 2777248 | EPI_ISL_ 2777385 |  |
| EPI_ISL_ 1261685 | EPI_ISL_ 2777249 | EPI_ISL_ 2777388 |  |
| EPI_ISL_ 1261690 | EPI_ISL_ 2777250 | EPI_ISL_ 2777397 |  |
| EPI_ISL_ 1261694 | EPI_ISL_ 2777251 | EPI_ISL_ 2777398 |  |
| EPI_ISL_ 2777236 | EPI_ISL_ 2777320 | EPI_ISL_ 2777399 |  |
| EPI_ISL_ 2777238 | EPI_ISL_ 2777363 | EPI_ISL_ 2777400 |  |
| EPI_ISL_ 2777248 | EPI_ISL_ 2777364 | EPI_ISL_ 2777401 |  |
| EPI_ISL_ 2777249 | EPI_ISL_ 2777373 | EPI_ISL_ 2777402 |  |
| EPI_ISL_ 2777250 | EPI_ISL_ 2777374 | EPI_ISL_ 2777403 |  |
| EPI_ISL_ 2777251 | EPI_ISL_ 2777375 | EPI_ISL_ 2777404 |  |
| EPI_ISL_ 2777320 | EPI_ISL_ 2777376 | EPI_ISL_ 2777405 |  |
| EPI_ISL_ 2777363 | EPI_ISL_ 2777377 | EPI_ISL_ 2777406 |  |
| EPI_ISL_ 2777364 | EPI_ISL_ 2777378 | EPI_ISL_ 2777407 |  |
| EPI_ISL_ 2777373 | EPI_ISL_ 2777380 | EPI_ISL_ 2777408 |  |
| EPI_ISL_ 2777374 | EPI_ISL_ 2777382 | EPI_ISL_ 2777410 |  |
| EPI_ISL_ 2777375 | EPI_ISL_ 2777383 | EPI_ISL_ 2777412 |  |
| EPI_ISL_ 2777376 | EPI_ISL_ 2777384 | EPI_ISL_ 2777413 |  |
| EPI_ISL_ 2777377 | EPI_ISL_ 2777385 | EPI_ISL_ 2777414 |  |
| EPI_ISL_ 2777378 | EPI_ISL_ 2777388 | EPI_ISL_ 2777415 |  |
| EPI_ISL_ 2777380 | EPI_ISL_ 2777397 | EPI_ISL_ 2777417 |  |
| EPI_ISL_ 2777382 | EPI_ISL_ 2777398 | EPI_ISL_ 2777418 |  |
| EPI_ISL_ 2777383 | EPI_ISL_ 2777399 | EPI_ISL_ 2777419 |  |
| EPI_ISL_ 2777384 | EPI_ISL_ 2777400 | EPI_ISL_ 2777454 |  |
| EPI_ISL_ 2777385 | EPI_ISL_ 2777401 | EPI_ISL_ 2777461 |  |
| EPI_ISL_ 2777388 | EPI_ISL_ 2777402 | EPI_ISL_ 2777462 |  |
| EPI_ISL_ 2777397 | EPI_ISL_ 2777403 | EPI_ISL_ 2777465 |  |
| EPI_ISL_ 2777398 | EPI_ISL_ 2777404 | EPI_ISL_ 2777466 |  |
| EPI_ISL_ 2777399 | EPI_ISL_ 2777405 | EPI_ISL_ 2777467 |  |
| EPI_ISL_ 2777400 | EPI_ISL_ 2777406 | EPI_ISL_ 2777469 |  |
| EPI_ISL_ 2777401 | EPI_ISL_ 2777407 | EPI_ISL_ 2777470 |  |
| EPI_ISL_ 2777402 | EPI_ISL_ 2777408 | EPI_ISL_ 2777472 |  |
| EPI_ISL_ 2777403 | EPI_ISL_ 2777409 | EPI_ISL_ 2777473 |  |
| EPI_ISL_ 2777404 | EPI_ISL_ 2777410 | EPI_ISL_ 2777474 |  |
| EPI_ISL_ 2777405 | EPI_ISL_ 2777412 | EPI_ISL_ 2777475 |  |
| EPI_ISL_ 2777406 | EPI_ISL_ 2777413 | EPI_ISL_ 2777477 |  |
| EPI_ISL_ 2777407 | EPI_ISL_ 2777414 | EPI_ISL_ 2777478 |  |
| EPI_ISL_ 2777408 | EPI_ISL_ 2777415 | EPI_ISL_ 2777479 |  |
| EPI_ISL_ 2777409 | EPI_ISL_ 2777416 | EPI_ISL_ 2777481 |  |
| EPI_ISL_ 2777410 | EPI_ISL_ 2777417 | EPI_ISL_ 2777482 |  |
| EPI_ISL_ 2777412 | EPI_ISL_ 2777418 | EPI_ISL_ 2777483 |  |
| EPI_ISL_ 2777413 | EPI_ISL_ 2777419 | EPI_ISL_ 2777485 |  |
| EPI_ISL_ 2777414 | EPI_ISL_ 2777420 | EPI_ISL_ 2777495 |  |
| EPI_ISL_ 2777415 | EPI_ISL_ 2777454 | EPI_ISL_ 2777498 |  |
| EPI_ISL_ 2777416 | EPI_ISL_ 2777460 | EPI_ISL_ 2777503 |  |
| EPI_ISL_ 2777417 | EPI_ISL_ 2777461 | EPI_ISL_ 2777507 |  |
| EPI_ISL_ 2777418 | EPI_ISL_ 2777462 | EPI_ISL_ 2777508 |  |
| EPI_ISL_ 2777419 | EPI_ISL_ 2777464 | EPI_ISL_ 2777539 |  |
| EPI_ISL_ 2777420 | EPI_ISL_ 2777466 | EPI_ISL_ 2777599 |  |
| EPI_ISL_ 2777454 | EPI_ISL_ 2777467 | EPI_ISL_ 2777698 |  |
| EPI_ISL_ 2777460 | EPI_ISL_ 2777468 | EPI_ISL_ 2777700 |  |
| EPI_ISL_ 2777461 | EPI_ISL_ 2777469 | EPI_ISL_ 2777701 |  |
| EPI_ISL_ 2777462 | EPI_ISL_ 2777470 | EPI_ISL_ 2777740 |  |
| EPI_ISL_ 2777464 | EPI_ISL_ 2777472 | EPI_ISL_ 2777986 |  |
| EPI_ISL_ 2777465 | EPI_ISL_ 2777473 | EPI_ISL_ 2777987 |  |
| EPI_ISL_ 2777466 | EPI_ISL_ 2777475 | EPI_ISL_ 2777993 |  |
| EPI_ISL_ 2777467 | EPI_ISL_ 2777477 | EPI_ISL_ 2777995 |  |
| EPI_ISL_ 2777468 | EPI_ISL_ 2777478 | EPI_ISL_ 2777996 |  |
| EPI_ISL_ 2777469 | EPI_ISL_ 2777481 | EPI_ISL_ 2777997 |  |
| EPI_ISL_ 2777470 | EPI_ISL_ 2777482 | EPI_ISL_ 2777998 |  |
| EPI_ISL_ 2777471 | EPI_ISL_ 2777495 | EPI_ISL_ 2777999 |  |
| EPI_ISL_ 2777472 | EPI_ISL_ 2777498 | EPI_ISL_ 2778000 |  |
| EPI_ISL_ 2777473 | EPI_ISL_ 2777499 | EPI_ISL_ 2778002 |  |
| EPI_ISL_ 2777474 | EPI_ISL_ 2777503 | EPI_ISL_ 2778005 |  |
| EPI_ISL_ 2777475 | EPI_ISL_ 2777508 | EPI_ISL_ 811149 |  |
| EPI_ISL_ 2777477 | EPI_ISL_ 2777516 | EPI_ISL_ 833136 |  |
| EPI_ISL_ 2777478 | EPI_ISL_ 2777539 | EPI_ISL_ 833139 |  |
| EPI_ISL_ 2777479 | EPI_ISL_ 2777698 | EPI_ISL_ 833140 |  |
| EPI_ISL_ 2777481 | EPI_ISL_ 2777701 | EPI_ISL_ 906071 |  |
| EPI_ISL_ 2777482 | EPI_ISL_ 2777740 | EPI_ISL_ 906077 |  |
| EPI_ISL_ 2777483 | EPI_ISL_ 2777986 | EPI_ISL_ 906081 |  |
| EPI_ISL_ 2777484 | EPI_ISL_ 2777987 | EPI_ISL_ 918500 |  |
| EPI_ISL_ 2777485 | EPI_ISL_ 2777995 | EPI_ISL_ 918502 |  |
| EPI_ISL_ 2777495 | EPI_ISL_ 2777996 | EPI_ISL_ 918503 |  |
| EPI_ISL_ 2777498 | EPI_ISL_ 2777997 | EPI_ISL_ 918506 |  |
| EPI_ISL_ 2777499 | EPI_ISL_ 2777998 | EPI_ISL_ 918508 |  |
| EPI_ISL_ 2777503 | EPI_ISL_ 2778002 | EPI_ISL_ 918509 |  |
| EPI_ISL_ 2777507 | EPI_ISL_ 2778005 | EPI_ISL_ 918511 |  |
| EPI_ISL_ 2777508 | EPI_ISL_ 811149 |  |  |
| EPI_ISL_ 2777509 | EPI_ISL_ 833136 |  |  |
| EPI_ISL_ 2777516 | EPI_ISL_ 833138 |  |  |
| EPI_ISL_ 2777539 | EPI_ISL_ 833139 |  |  |
| EPI_ISL_ 2777599 | EPI_ISL_ 833140 |  |  |
| EPI_ISL_ 2777698 | EPI_ISL_ 906071 |  |  |
| EPI_ISL_ 2777700 | EPI_ISL_ 906080 |  |  |
| EPI_ISL_ 2777701 | EPI_ISL_ 906081 |  |  |
| EPI_ISL_ 2777740 | EPI_ISL_ 918500 |  |  |
| EPI_ISL_ 2777986 | EPI_ISL_ 918501 |  |  |
| EPI_ISL_ 2777987 | EPI_ISL_ 918502 |  |  |
| EPI_ISL_ 2777993 | EPI_ISL_ 918503 |  |  |
| EPI_ISL_ 2777995 | EPI_ISL_ 918505 |  |  |
| EPI_ISL_ 2777996 | EPI_ISL_ 918506 |  |  |
| EPI_ISL_ 2777997 | EPI_ISL_ 918507 |  |  |
| EPI_ISL_ 2777998 | EPI_ISL_ 918508 |  |  |
| EPI_ISL_ 2777999 | EPI_ISL_ 918510 |  |  |
| EPI_ISL_ 2778000 | EPI_ISL_ 918511 |  |  |
| EPI_ISL_ 2778002 |  |  |  |
| EPI_ISL_ 2778004 |  |  |  |
| EPI_ISL_ 2778005 |  |  |  |
| EPI_ISL_ 811149 |  |  |  |
| EPI_ISL_ 833136 |  |  |  |
| EPI_ISL_ 833138 |  |  |  |
| EPI_ISL_ 833139 |  |  |  |
| EPI_ISL_ 833140 |  |  |  |
| EPI_ISL_ 906071 |  |  |  |
| EPI_ISL_ 906075 |  |  |  |
| EPI_ISL_ 906076 |  |  |  |
| EPI_ISL_ 906077 |  |  |  |
| EPI_ISL_ 906080 |  |  |  |
| EPI_ISL_ 906081 |  |  |  |
| EPI_ISL_ 918499 |  |  |  |
| EPI_ISL_ 918500 |  |  |  |
| EPI_ISL_ 918501 |  |  |  |
| EPI_ISL_ 918502 |  |  |  |
| EPI_ISL_ 918503 |  |  |  |
| EPI_ISL_ 918504 |  |  |  |
| EPI_ISL_ 918505 |  |  |  |
| EPI_ISL_ 918506 |  |  |  |
| EPI_ISL_ 918507 |  |  |  |
| EPI_ISL_ 918508 |  |  |  |
| EPI_ISL_ 918509 |  |  |  |
| EPI_ISL_ 918510 |  |  |  |
| EPI_ISL_ 918511 |  |  |  |

